# Amyloid and Tau Neuropathologic Stage, Coexisting Brain Disease, and Cognitive Status in a Population-Based Autopsy Cohort

**DOI:** 10.64898/2026.10.20.26365945

**Authors:** Felipe L. Pereira, Renata E. P. Leite, Vitor Paes, Caroline M. Silva, Raul R. Ururahy, Filipe O. C. Moraes, Ines L. Padrao, Carlos Pasqualucci, Roberta D. Rodrigues, Michel Naslavsky, Renata E. L. Ferretti-Rebustini, Ricardo Nitrini, Eduardo Ferrioli, Kristine Yaffe, Claudia K. Suemoto, Lea T. Grinberg

**Author notes:** Equal contributions (Claudia K. Suemoto, Lea T. Grinberg). Corresponding Author: Lea T. Grinberg, MD, PhD, Department of Laboratory Medicine and Pathology, Mayo Clinic, 4500 San Pablo Road, Jacksonville, FL 32224.

## Abstract

**Importance:** Interpreting an amyloid or tau biomarker result requires knowing how often the target pathology is present in the tested population, whether a single marker characterizes the joint amyloid-tau state, and whether the detected pathology explains the patient symptoms. Population-based autopsy data addressing these questions are scarce, and recent population-scale prevalence estimates rest on biomarkers rather than ground-truth tissue examination.

**Objective:** To estimate stage-resolved, age- and sex-specific frequencies of amyloid and tau neuropathology in an unselected, admixed, autopsy population and to evaluate their clinical expression in relation to coexisting neuropathology.

**Design Setting and Participants:** Cross-sectional clinicopathologic study of decedents accrued from 2004 through 2025 at the Biobank for Aging Studies of the University of Sao Paulo, sampled through the centralized Sao Paulo municipal autopsy service, which performs approximately 15 000 autopsies annually and verifies about 16% of deaths in the city. Eligible decedents were aged 18 years or older with complete amyloid, tau, Clinical Dementia Rating (CDR), and coexisting pathology assessments.

**Exposures:** Age, sex, Thal amyloid phase, Braak neurofibrillary tangle stage, Consortium to Establish a Registry for Alzheimer Disease (CERAD) neuritic plaque score, and contributing coexisting neuropathology.

**Main Outcomes and Measures:** The complete Thal phase by Braak stage distribution; age- and sex-specific frequency of pathology in the biomarker-detectable range, prespecified as Thal phase 3 to 5 together with Braak stage IV to VI; and associations with CDR and CDR Sum of Boxes (CDR-SB).

**Results:** Among 2711 decedents (mean [SD] age, 73.4 [14.4] years; range, 23 to 111 years; 1340 [49.4%] female), 531 (19.6%) met both thresholds, 270 (10.0%) the amyloid threshold only, 207 (7.6%) the tau threshold only, and 1703 (62.8%) neither. Frequency of the combined state rose from 0% at ages 23 to 50 years to 5.5% at 61 to 70, 14.5% at 71 to 80, 36.5% at 81 to 90, and 45.9% at 91 to 111 years. The proportion with CDR greater than 0 increased monotonically across Braak stages, from 88 of 541 (16.3%) at stage 0 to 155 of 179 (86.6%) at stage VI. When the combined amyloid-tau state was cross-classified with contributing coexisting pathology, CDR greater than 0 was present in 225 of 1467 participants (15.3%) with neither, 204 of 349 (58.5%) with coexisting pathology alone, 216 of 339 (63.7%) with combined amyloid-tau pathology alone, and 165 of 192 (85.9%) with both. In a multivariable model of CDR-SB, contributing coexisting pathology had the largest unique adjusted contribution (partial *R*^2^, 0.088; 95% CI, 0.064-0.114), and Braak stage had the largest share of total explained variance (Lindeman, Merenda, and Gold contribution, 0.115).

**Conclusions and Relevance:** Amyloid and tau pathology within the range detected by current biomarkers was uncommon before age 70 and discordant in 17.6% of decedents. Contributing coexisting brain disease, which amyloid and tau biomarkers do not detect, accompanied cognitive impairment as often as advanced Alzheimer disease pathology did. These tissue-based frequencies can inform the interpretation of positive and negative biomarker results.

**Key Points:** *Question:* Among people with amyloid and tau neuropathology at levels expected to be detectable by current biomarkers, how often is cognitive impairment present, and how does coexisting brain disease affect this relationship?

*Findings:* In this cross-sectional study of 2711 autopsies from a population-based cohort aged 23 to 111 years, 19.6% had neuropathology meeting both amyloid and tau thresholds expected to be detectable by current biomarkers, rising from 0% before age 51 to 45.9% after age 90. Contributing coexisting brain disease alone was accompanied by impairment in 58.5% of participants, nearly as often as amyloid and tau pathology alone (63.7%).

*Meaning:* Age-specific neuropathologic frequencies provide an autopsy-derived context for interpreting the predictive value of Alzheimer disease biomarkers, while coexisting brain disease was a major contributor to cognitive impairment beyond amyloid and tau pathology.

## Introduction

Alzheimer disease (AD) is defined biologically by amyloid β plaques and tau neurofibrillary tangles ^1,2^. Neuropathologic assessment integrates amyloid plaque distribution, neuritic plaque burden, and neurofibrillary tangle distribution, commonly summarized by Thal amyloid phase, Consortium to Establish a Registry for AD (CERAD) score, and Braak stage ^3^. Positron Emission Tomography (PET), cerebrospinal fluid, and blood biomarkers of amyloid and tau increasingly guide diagnosis, prognosis, trial enrollment, and treatment. Their interpretation depends not only on assay performance but also on the frequency and clinical relevance of the underlying pathology in the population tested.

Biomarker interpretation raises 3 questions. First, does a positive result indicate that the target pathology is present? Positive predictive value, the probability that the target pathology is present given a positive test result, depends on sensitivity, specificity, and the frequency of the pathology. Because that frequency varies with age, cognitive status, apolipoprotein E (APOE) ε4 status, and referral setting, the same test result can yield different posttest probabilities ^4–6^.

Second, is a single AD biomarker sufficient to characterize the relevant pathologic state? Amyloid and tau are related but not interchangeable. Tau distribution and neuritic plaque burden are more closely associated with cognitive and functional impairment than amyloid distribution alone ^1,3,7^. Discordant amyloid and tau states may therefore limit inference from a single marker. Third, does the detected AD pathology explain the patient symptoms and inform treatment expectations? AD pathology may be clinically silent, the primary contributor to impairment, or one of several contributors. Vascular disease, Lewy body disease, hippocampal sclerosis, transactive response DNA-binding protein 43 (TDP-43) pathology, and other age-related lesions can contribute to the same presentation but are incompletely captured or not measured by current amyloid and tau biomarkers ^8–11^.

The rapid adoption of blood biomarkers makes these interpretation challenges more urgent. Plasma phosphorylated tau 217 (p-tau217) is increasingly used for screening and triage ^12^, and community studies now use it to estimate the frequency of biomarker-defined AD biology ^13^. In a Norwegian population-based study of 11 486 plasma samples, the estimated frequency of AD neuropathologic change rose from under 8% before age 70 years to 65.2% after age 90 years, and approximately 11% of participants aged 70 years or older met current eligibility criteria for anti-amyloid therapy. The investigators noted that their estimates are potentially circular because the frequency used to anchor predictive values was derived from the same assay under evaluation, and that they could not examine whether coexisting pathology contributed to cognition ^13^. Blood biomarker performance is often evaluated against PET or cerebrospinal fluid rather than autopsy. Because these comparators are also proxies for tissue pathology, concordance among biomarkers does not directly define the complete neuropathologic state ^14,15^.

Autopsy remains the reference standard for direct characterization of amyloid, tau, neuritic plaques, and contributing coexisting brain diseases, and it is the only method that can establish how frequently each pathologic stage occurs in an unselected population and how often coexisting disease accompanies it; no in vivo measurement can substitute because the quantity of interest is the tissue state itself. Yet population-based autopsy data are limited, particularly for adults younger than 80 years and populations underrepresented in brain donor and biomarker cohorts ^16,17^. Existing population-based autopsy series, including the Cognitive Function and Aging Study ^18,19^, the Vantaa 85+ study ^20^, the Hisayama study ^21^, the Honolulu-Asia Aging Study ^22^, and the 90+ Study ^23^, are substantially smaller and almost entirely restricted to the oldest old, so they cannot resolve stage-specific frequency below age 80 years or in racially diverse populations. This limits tissue-based frequency estimates and often makes another biomarker, rather than neuropathology, the reference for estimating predictive value.

We studied 2711 decedents from the Biobank for Aging Studies at the University of Sao Paulo, a population-based cohort sampled through a centralized municipal autopsy service. We characterized the full Thal by Braak matrix and CERAD neuritic plaque burden, estimated age- and sex-specific frequency of biomarker-detectable amyloid and tau tissue states, and evaluated their associations with cognitive and functional status and contributing coexisting pathology. Our objective was to provide autopsy-derived estimates for target tissue states, characterize concordant and discordant amyloid and tau states, and examine how often these states were accompanied by impairment.

## Methods

### Study Design and Participants

This cross-sectional study included participants accrued from 2004 through 2025 in the Biobank for Aging Studies at the University of Sao Paulo (BAS-USP), an ongoing brain bank linked to the centralized Sao Paulo Autopsy Service. The service receives nontraumatic deaths requiring medical verification of the cause or sequence leading to death and performs approximately 15 000 autopsies annually. In a prior assessment covering 2004 through 2014, 16% of city deaths underwent autopsy at the service (this rate is among the highest reported for any medical autopsy service worldwide), and BAS-USP participants had age and sex distributions similar to citywide deaths ^17,24^. To assess the demographic representativeness of this specific study cohort relative to the citywide mortality population, we generated 10 000 bootstrap samples from the citywide mortality database, each matched to the study cohort size. Comparisons showed no evidence of differences in sex proportions after false discovery rate (FDR) adjustment (χ² test, P = .90) or in age distributions among individuals older than 50 years when stratified by sex (females, P = .68; males, P = .78). The density of age at death to both sexes is presented in **eFigure 1**. Written consent was obtained from the next of kin, and the study was approved by the University of São Paulo Board of Committee Review ^25^. Recruitment, consent, and cohort procedures are described in the eMethods ^26,27^.

Of 2716 available participants, 3 were excluded for missing Clinical Dementia Rating (CDR) or CDR Sum of Boxes (CDR-SB) data, and 2 for missing Thal phase or Braak stage, leaving 2711 participants with complete primary clinicopathologic data. Race was available for 2061 participants, education for 2066, and APOE genotyping for 1518. The primary frequency estimate was the distribution at death of amyloid and tau neuropathologic states among eligible decedents in the municipal autopsy service source population.

### Clinical Assessment

A trained gerontologist interviewed the most knowledgeable informant using a standardized protocol referring to cognitive and functional status during the final 3 months of life ^17,25^. Global CDR scores ranged from 0 to 3 ^28^; CDR greater than 0 identified cognitive or functional impairment, and CDR-SB was analyzed continuously. The informant assessment has been validated against memory clinic consensus diagnosis ^29^. Age and sex were obtained from official or autopsy service records. Education and race were reported by the informant. Race response options followed the Brazilian Institute of Geography and Statistics categories of White, Black, Pardo, Indigenous, and Asian; Black and Pardo responses were combined as Black/Brown for analysis, consistent with prior BAS-USP studies, and no participant was classified as Indigenous. APOE genotyping and further details on demographic ascertainment are described in the eMethods ^26,27^.

### Neuropathologic Assessment

Neuropathologic evaluation used systematic regional sampling, hematoxylin and eosin staining, and immunohistochemistry for amyloid β, phosphorylated tau, TDP-43, and α-synuclein. Experienced neuropathologists assigned diagnoses and stages without knowledge of the clinical assessment ^17,25–27^. The Thal phase measured amyloid plaque distribution, the Braak stage measured neurofibrillary tangle distribution, and the CERAD score graded neocortical neuritic plaque density ^3,30–32^.

No participant had antemortem biomarker measurements available for this study. Prespecified thresholds defined biomarker-detectable amyloid pathology as Thal phase 3 to 5, biomarker-detectable tau pathology as Braak stage IV to VI, and combined pathology as meeting both thresholds. We use “biomarker-detectable” to denote stages at which current amyloid and tau assays are expected to yield positive results; the correspondence is inferred from published autopsy-validation studies and was not measured here. The Thal phase 3 was selected because it represents amyloid distribution generally associated with a detectable amyloid PET signal ^14,33^, and Braak stage IV because quantitative regional flortaucipir approaches detect intermediate tau burden at approximately this stage, whereas standard visual interpretation corresponds more closely to Braak stages V to VI ^34–36^. Contributing coexisting pathology (yes or no) comprised cerebrovascular disease, limbic-predominant age-related TDP-43 encephalopathy neuropathologic change stages 2 or 3, neocortical Lewy body pathology, hippocampal sclerosis, and/or primary frontotemporal lobar degeneration with sufficient burden or distribution to contribute to impairment ^8–10,37^. Detailed operational definitions are provided in the eMethods.

### Statistical Analysis

The primary descriptive representation was the complete Thal phase by Braak stage matrix, with the CERAD score analyzed separately. Summary analyses classified participants into 4 mutually exclusive states: neither threshold, amyloid-only, tau-only, or both thresholds. Age strata were 23 to 50, 51 to 60, 61 to 70, 71 to 80, 81 to 90, and 91 to 111 years. All tests were 2-sided with alpha = 0.05. Analyses used R version 4.6.1 and its packages as described in eMethods.

Clinical attribution was evaluated by comparing the proportion with CDR greater than 0 across tissue states and after cross-classification with contributing coexisting pathology. Absolute differences in the principal proportions were reported with 95% CIs. Two multivariable linear regressions related to CDR-SB were performed: first, *m0* accounting to Thal phase, Braak stage, CERAD score, contributing coexisting pathology, age, sex, APOE ε4 status, and education, using participants with complete data for all model variables (n = 1518, 56.0% of the cohort), and second *m1* (n = 2711, 100% of the cohort) excluding education and APOE ε4 status due to the missing data and lower or null participation in *m0*. Partial *R2* was used to quantify unique adjusted contributions, and the Lindeman, Merenda, and Gold (LMG) method was used to explain the total variance. We use partial *R*^2^ and LMG together because partial *R*^2^ quantifies each predictor’s unique contribution after adjustment for all others, whereas LMG distributes both unique and shared explained variance, which is especially important when predictors are correlated. Fractional logit sensitivity analyses, bootstrap procedures, cross-validation, missing-data handling, model diagnostics, and candidate models are detailed in the eMethods and eResults.

In an exploratory analysis, we combined the observed tissue frequencies with externally derived sensitivity and specificity ranges for plasma p-tau217 to estimate age-specific positive and negative predictive values by the Bayes formula, reporting both unpenalized and optimism-corrected values. Sensitivity and specificity were taken from an antemortem-postmortem cohort in which plasma was collected 0.02 to 2.9 years before death and the reference standard was neuropathologic diagnosis (sensitivity 94%, specificity 94% for Braak stage V to VI with moderate-to-frequent neuritic plaques; sensitivity 94%, specificity 79% for the broader intermediate-to-high category) ^38^; the range derived against amyloid PET and cerebrospinal fluid used in prior population estimates ^12^ is shown for comparison. Unlike predictive values anchored to biomarker-defined frequency, these estimates use a tissue-defined prior.

## Results

### Cohort Characteristics

The analytic cohort included 2711 participants (mean [SD] age at death, 73.4 [14.4] years; range, 23 to 111 years); 1340 (49.4%) were female, 1787 (65.9%) had CDR 0, 924 (34.1%) had CDR greater than 0, and 541 (20.0%) had contributing coexisting pathology (**Figure 1** and **Table 1**). Among participants with available data, 753 of 2061 (36.5%) were black or brown, and 450 of 1518 (29.6%) were APOE ε4 carriers.

**Figure 1.**
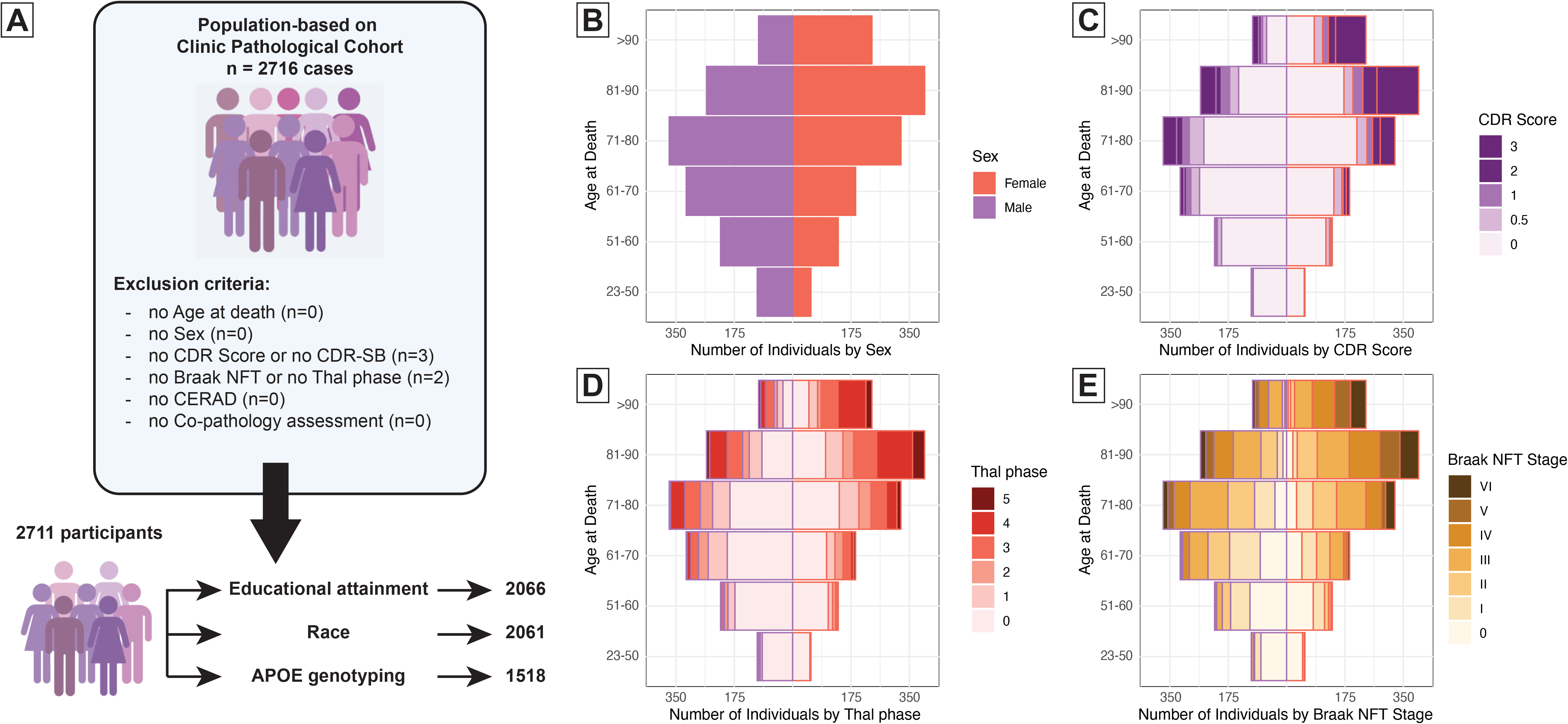
Cohort Derivation and Age-at-Death Distributions. **A**, Cohort derivation from 2716 participants screened and enrolled during recruitment periods. Exclusions were missing Clinical Dementia Rating (CDR) or CDR Sum of Boxes (CDR-SB) data (n = 3) and missing Braak neurofibrillary tangle (NFT) stage or Thal amyloid phase (n = 2). No participants were missing age at death, sex, Consortium to Establish a Registry for Alzheimer’s Disease (CERAD) neuritic plaque score, or assessment of contributing coexisting pathology. The final analytic cohort included 2711 participants. Race was available for 2061 participants, education for 2066, and APOE genotyping for 1518. **B through E**, Age-at-death distributions by sex, CDR score, Thal amyloid phase, and Braak neurofibrillary tangle stage. Abbreviations: APOE, apolipoprotein E; CDR, Clinical Dementia Rating; CDR-SB, CDR Sum of Boxes; NFT, neurofibrillary tangle; CERAD, Consortium to Establish a Registry for Alzheimer Disease.

**Table 1.**
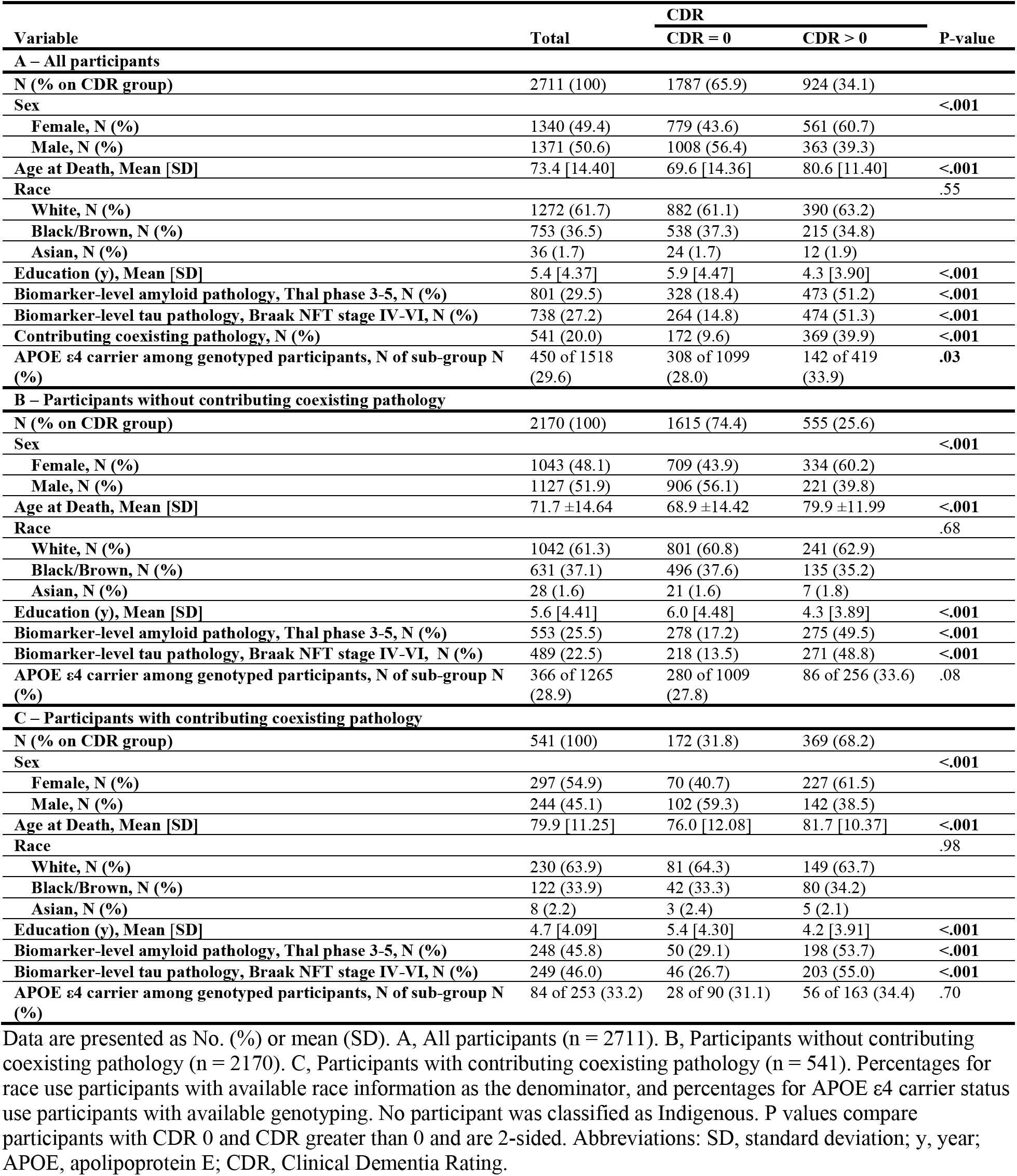
Demographic, Clinical, and Neuropathologic Characteristics by Clinical Dementia Rating and Contributing Coexisting Pathology.

### Frequency of Biomarker-Detectable Amyloid and Tau Pathology

Biomarker-detectable amyloid pathology was present in 801 participants (29.5%), biomarker-detectable tau pathology in 738 (27.2%), and both thresholds in 531 (19.6%) (**Figure 2A**). The full matrix retained lower-burden, concordant, and discordant tissue states (**eTable 1**), and the CERAD score was summarized separately (**eTable 2**). Combined pathology was present in 0% at ages 23 to 50 years, 2.0% at ages 51 to 60 years, 5.5% at ages 61 to 70 years, 14.5% at ages 71 to 80 years, 36.5% at ages 81 to 90 years, and 45.9% at ages 91 to 111 years (**eFigure 2**). Among participants who met the amyloid threshold, 66.3% also met the tau threshold; among those who met the tau threshold, 72.0% also met the amyloid threshold.

**Figure 2.**
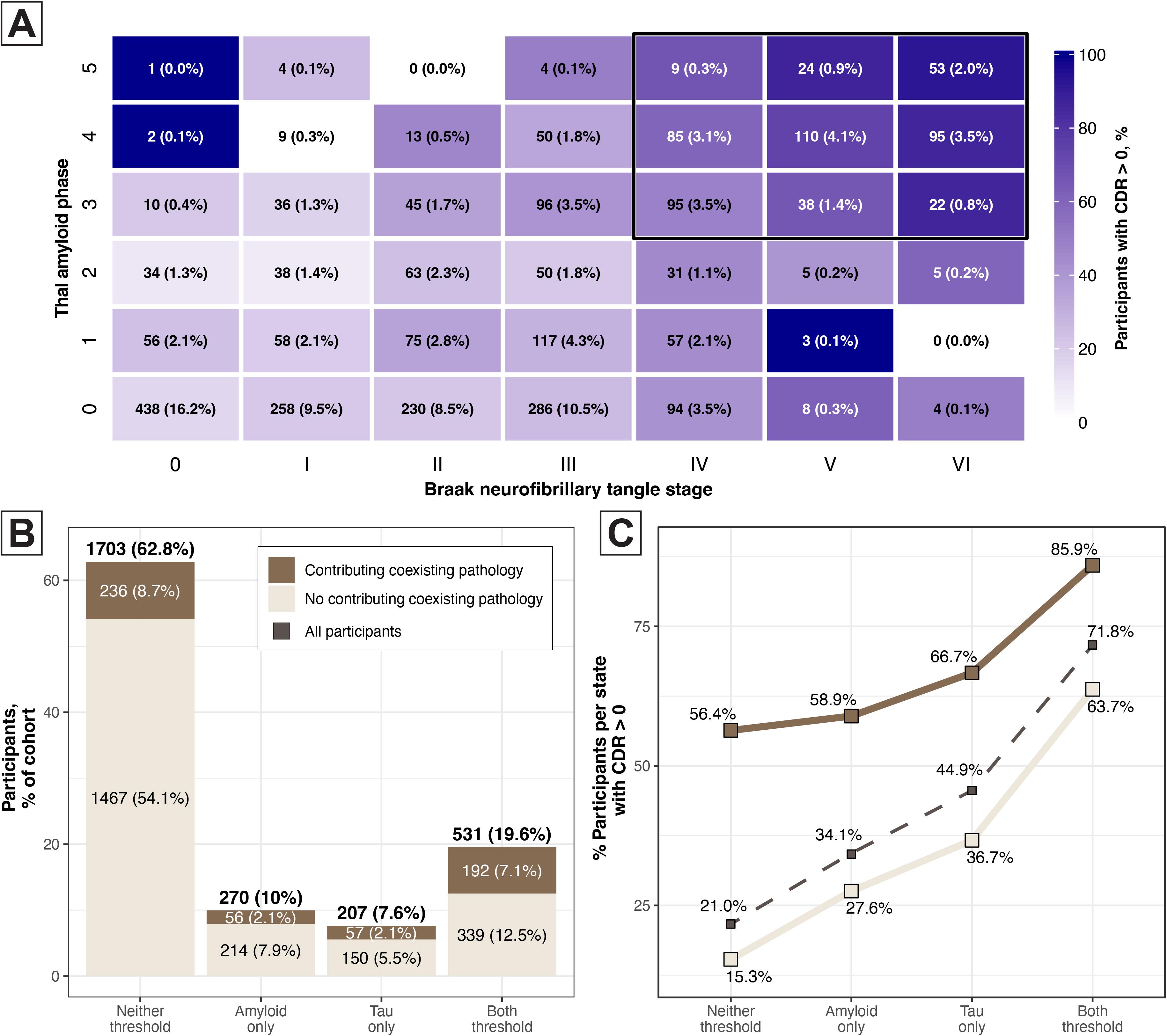
Joint Thal Amyloid Phase and Braak Neurofibrillary Tangle Stage and Its Clinical Expression (n = 2711). **A**, Joint distribution of Thal amyloid phase and Braak neurofibrillary tangle stage. Each cell shows the number of participants, the cohort percentage in parentheses. Shading indicates the percentage with CDR greater than 0 in each cell. The outlined region indicates combined biomarker-detectable amyloid and tau pathology, defined as Thal phase 3 to 5 together with Braak stage IV to VI. **B and C**, The same participants were classified into 4 mutually exclusive states. **B**, Panel shows the number and percentage of participants in each state, divided into participants with and without contributing coexisting pathology; above each bar is the state total. **C**, Panel shows the percentage of participants with CDR greater than 0 in each state with (solid dark brown line) and without (solid light brown line) contributing coexisting pathology, as well as all participants (dashed gray line). Abbreviations: %, percentage; CDR, Clinical Dementia Rating.

Frequency of combined biomarker-detectable pathologies did not differ significantly between female and male participants at ages 70 years or younger and was higher among female participants at older ages; at ages 91 to 111 years, frequency was 54.2% in female participants and 26.5% in male participants (**eFigure 3**). Because female participants died at older ages, the crude difference between women and men across the whole cohort (28.1% vs 11.3%) largely reflects age; age-stratified estimates are the interpretable comparison (**eFigure 3 and eTables 3 and 4**). An exploratory comparison by race group is shown in **eFigure 4**.

### Clinical Attribution and Contributing Coexisting Pathology

The proportion of participants with CDR greater than 0 increased monotonically across Braak stage, from 88 of 541 (16.3%) at stage 0 to 74 of 403 (18.4%) at stage I, 112 of 426 (26.3%) at stage II, 176 of 603 (29.2%) at stage III, 181 of 371 (48.8%) at stage IV, 138 of 188 (73.4%) at stage V, and 155 of 179 (86.6%) at stage VI (**eTable 1**).

The proportion with CDR greater than 0 also increased across the 4 mutually exclusive amyloid and tau states: 358 of 1703 participants (21.0%) meeting neither threshold, 92 of 270 (34.1%) meeting the amyloid threshold only, 93 of 207 (44.9%) meeting the tau threshold only, and 381 of 531 (71.8%) meeting both (**Figure 2B-C**). Overall, CDR greater than 0 was present in 71.8% of participants meeting both thresholds and 24.9% summing the other 3 states, an absolute difference of 46.8 percentage points (95% CI, 42.5 to 50.9; *P* < .001) (**Figure 2B-C**).

Restricting to the 2170 participants without contributing coexisting pathology, the corresponding proportions were 15.3%, 27.6%, 36.7%, and 63.7%, indicating that the gradient across tissue states persisted after coexisting disease was excluded (**Figure 2B-C**). The complete distribution among the 2170 participants without contributing coexisting pathology is shown in **eTable 5 and eFigure 5**.

### Factors Associated with Functional Cognitive Impairment Severity

In the primary adjusted model *m0* (n = 1518), education was inversely associated with CDR-SB but had a small unique contribution (partial *R*^2^, 0.007), whereas APOE ε4 status was not independently associated (P = .26; partial *R*^2^, 0.001) (**Figure 3A** and **eFigure 6**). Given their substantial missingness and limited contribution, both variables were omitted from the secondary adjusted model *m1*, which included all 2711 participants. In *m1*, Braak stage VI was associated with a 6.13-point higher CDR-SB score (95% CI, 4.932 to 7.324), contributing coexisting pathology with a 4.03-point (95% CI, 3.538 to 4.517), Braak stage V with 3.34-point (95% CI, 2.212 to 4.466), CERAD 3 with 2.38-point (95% CI, 1.212 to 3.558), and Thal phase 5 with 2.19-point (95% CI, 0.735 to 3.642). Female sex and older age at death had smaller adjusted associations. Related work from this biobank found that the association between female sex and neuritic plaque burden was attenuated after adjustment for Braak stage, highlighting the interdependence of sex-specific amyloid and tau findings ^39^. The complete categories and variables association can is found in **eTable 6**.

**Figure 3.**
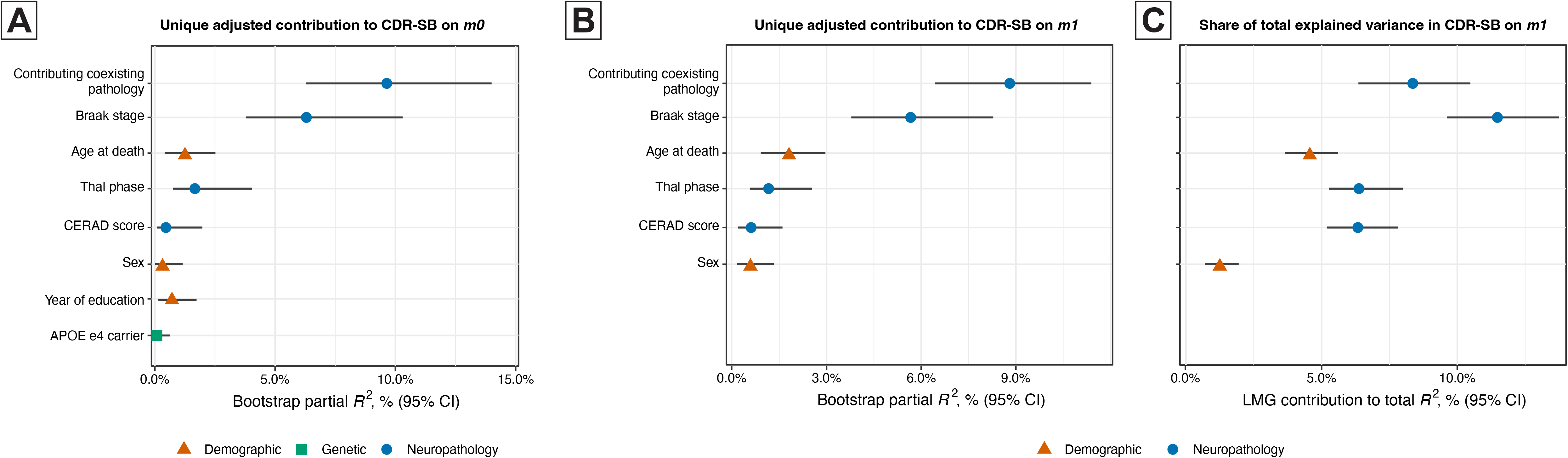
Contributing Coexisting Pathology and Relative Importance of Predictors of Cognitive and Functional Impairment. A and B, Term-level partial *R*^2^ with percentile bootstrap 95% CIs, representing the unique adjusted contribution of each predictor after accounting for all remaining variables. C, Lindeman, Merenda, and Gold contributions to total R2 with bootstrap 95% CIs. In panels A and B, Thal phase, Braak stage, and CERAD neuritic plaque score were modeled as categorical factors; Marker color indicates the neuropathologic, demographic, and genetic domains. Panel A is based on 1518 participants with complete data for all model variables in model *m0*. Panels B and C are based on all participants (n = 2711) in model *m1*. Abbreviations: APOE, apolipoprotein E; CDR-SB, Clinical Dementia Rating Sum of Boxes; CERAD, Consortium to Establish a Registry for Alzheimer’s Disease; CI, confidence interval.

In secondary robust model variation, contributing coexisting pathology had the largest unique contribution to CDR-SB variation (**Figure 3B**, partial *R2*, 0.089; 95% CI, 0.064-0.114; ranked first in 95.4% of bootstrap samples), whereas Braak stage had the largest contribution to total explained variance (**Figure 3C**, LMG contribution, 0.115; ranked first in 97.5% of bootstrap samples). CERAD neuritic plaque score was supported by global heteroskedasticity-consistent tests. Fractional logit sensitivity analyses yielded similar term-level conclusions and kept all predictions within the valid CDR-SB range; detailed diagnostics and cross-validation are shown in **eFigure 7**.

### Exploratory Predictive Values

Combining the observed tissue frequencies with externally ^38^ derived reported sensitivity of 94% and specificity of 94% ranges using moderate-to-frequent neuritic plaques for plasma p-tau217, the estimated positive predictive value rose with age from 47.7% at ages 61 to 70 years to 72.7% at ages 71 to 80 years and 93.0% at ages 91 to 111 years, while the negative predictive value fell from 99.6% to 94.9%. The positive predictive values dropped to 21.4%, 44.1%, and 78.9% for the same age strata when a sensitivity of 94% and specificity of 79% were used, considering the broader intermediate-to-high neuritic plaques category from the same report ^38^ (**Figure 4**). The positive predictive value reached 50% once the frequency of combined target pathology exceeded ∼6% in the moderate-to-frequent estimation (Se 94% / Sp 94%) or ∼18% in the intermediate-to-high estimation (Se 94% / Sp 79%). Values for all age strata in Se 0.850-0.982 / Sp 0.745-0.986, including optimism-corrected estimates, are given in **eTable 7**. Because the prior is tissue-derived, these estimates are not subject to the circularity that arises when frequency is estimated from the same biomarker being evaluated.

**Figure 4.**
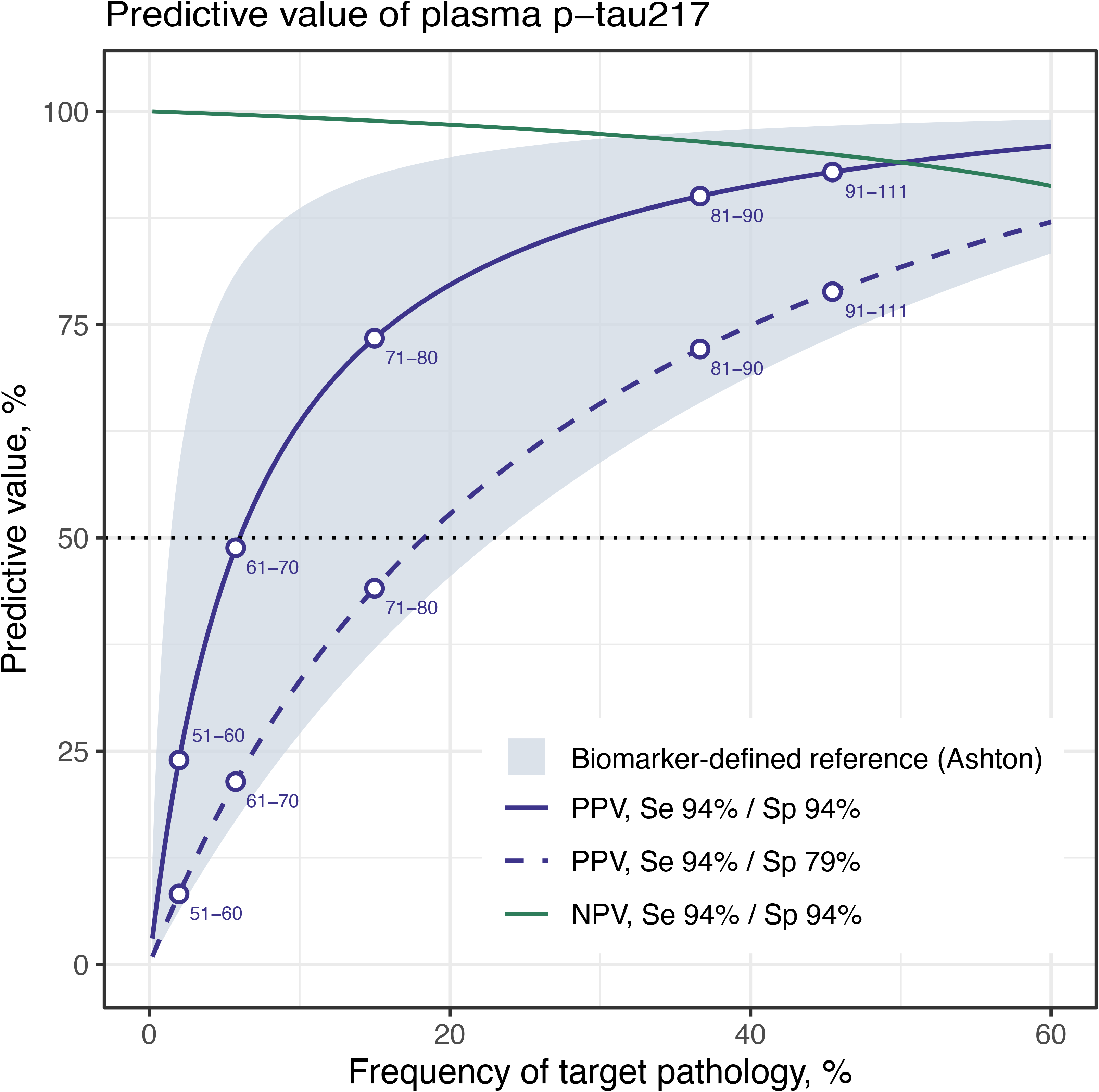
Estimated Predictive Values for Plasma Phosphorylated Tau 217. Estimated positive and negative predictive values for a plasma phosphorylated tau 217 assay, plotted as continuous functions of the frequency of the target pathology. Solid and dashed lines show positive predictive value at 2 operating points derived from an antemortem-postmortem cohort; the third line shows negative predictive value at the first of these. The shaded band shows the range of positive predictive values obtained from the sensitivity and specificity ranges derived from amyloid positron emission tomography and cerebrospinal fluid used in population biomarker studies. Open circles mark the age-specific frequencies derived from the combined biomarker-detectable-level pathology of our study. The dotted horizontal line marks a predictive value of 50%. These estimates are exploratory and assume that externally derived sensitivity and specificity transfer to this population and that frequency at death approximates the pretest probability in the population being tested.

## Discussion

In this population-based autopsy study, tissue findings informed three questions central to AD biomarker interpretation: how frequently the intended tissue state is present, whether one marker adequately characterizes the joint amyloid-tau state, and whether the detected pathology explains the impairment. Biomarker-detectable combined amyloid and tau pathology increased sharply with age, 17.6% of participants met only 1 of the 2 thresholds, and the combined state was strongly but incompletely associated with impairment. Contributing coexisting pathology was associated with additional impairment and outranked every AD measure in its unique adjusted contribution to impairment severity, despite being the one category of pathology that current amyloid and tau biomarkers cannot detect. Because antemortem biomarkers were unavailable, these categories represent tissue states inferred to fall within the detection range of current assays, rather than observed biomarker results.

Age-specific tissue frequency provides a pretest probability for interpreting positive and negative results, and it can be obtained only at autopsy because the quantity of interest is the tissue state (ground truth) rather than a correlate of it. Combined pathology was present in 5.5% of participants aged 61 to 70 years and 14.5% of those aged 71 to 80 years, age groups that include many patients evaluated for cognitive symptoms and enrolled in trials or in anti-amyloid treatment ^40,41^. Current blood biomarker guidance emphasizes use within a comprehensive clinical evaluation and explicit consideration of pretest probability ^42^. In cognitively unimpaired cohorts, the positive predictive value of plasma p-tau217 increased from 38% before age 60 years to 93% at age 80 years or older, while the negative predictive value decreased from 91% to 66% ^43^.

A Norwegian population-based study of 11 486 plasma samples found a similar age gradient using plasma p-tau217, but biomarker-defined frequency exceeded tissue-defined frequency at every comparable age: approximately 22% versus 14.5% at ages 71 to 80 years, approximately 49% versus 36.5% at ages 81 to 90 years, and 65.2% versus 45.9% after age 90 years ^13^. The 2 sets of estimates are not directly comparable because the populations, thresholds, and pathologic constructs differ, and because our estimates describe frequency at death in an autopsy source population, whereas theirs describe frequency among living survey participants and were weighted for nonparticipation. Even allowing for these differences, biomarker-defined frequency ran 34-52% higher than tissue-defined frequency across the age range. This matters for planning because treatment-eligibility projections scale with the assumed frequency: that study estimated that about 11% of people aged 70 years or older would meet current criteria for anti-amyloid therapy. Biomarker-defined and tissue-defined frequency are related but not interchangeable, and predictive values anchored to the former inherit the circularity the authors acknowledged.

The full Thal by Braak matrix also showed why one binary result may incompletely characterize the pathological state. Amyloid-only and tau-only (biomarker-detectable) states accounted for 10.0% and 7.6% of participants, respectively, and many additional participants had lower-burden states below the prespecified thresholds. Autopsy-linked studies document discordance between amyloid PET, cerebrospinal fluid, and tissue pathology; reduced tau PET sensitivity for lower-stage pathology and primary age-related tauopathy; and clinically relevant AD contributing coexisting pathology in syndromes usually associated with non-AD disease ^44–47^. Biomarkers also vary in their ability to detect early stages of amyloid and tau ^14,48^. These findings support joint amyloid and tau assessments, intermediate-range results, and additional modalities when the biomarker results and clinical phenotypes do not align. Because the matrix is reported in full (**Figure 2A**), frequencies can be recomputed for any future threshold pair as assay sensitivity changes, which a biomarker-defined estimate cannot provide. The adjusted associations further distinguished among related tissue dimensions: Braak stage was consistently associated with impairment severity, whereas Thal phase was only nonmonotonically associated, and the CERAD association depended on the model specification.

Symptom attribution remained incomplete even when both thresholds were met. Impairment (CDR > 0) was identified in 71.8% of participants meeting both thresholds, leaving 28.2% without identified impairment; conversely, 24.9% of participants in the other tissue states had impairment. Contributing coexisting pathology increased the frequency of impairment among participants meeting both thresholds from 63.7% to 85.9%, and among participants who did not meet both amyloid and tau thresholds, coexisting pathology alone was accompanied by impairment in 58.5%, nearly as often as combined amyloid and tau pathology alone (63.7%).

The contributing processes included vascular disease, limbic-predominant age-related TDP-43 encephalopathy, neocortical Lewy body pathology, hippocampal sclerosis, and frontotemporal lobar degeneration, all of which are established contributors to late-life cognitive impairment ^8–10,37^. In a related analysis from the same biobank, advanced Lewy body pathology was associated with worse cognition and psychosis-related symptoms in a mixed-pathology context. The present analysis places selected coexisting pathologies within the joint distribution of amyloid and tau stages ^11^. Except for cerebrovascular changes and severe Lewy body disease, these comorbidities are not detectable by current biomarkers, and the recent population-scale biomarker literature has explicitly left their contribution to cognition unexamined ^13^. This distinction is clinically relevant as anti-amyloid treatment expands in biomarker-confirmed early AD ^40,41^. The present study did not evaluate therapy, but it identifies tissue sources of heterogeneity among patients with similar

AD biomarker profiles. Relative-importance analyses were consistent with this interpretation: contributing coexisting pathology had the strongest unique association with CDR-SB, while Braak stage accounted for the largest overall share of explained variance. These estimates describe adjusted associations rather than causal effects.

The cohort adds information that is not routinely available in specialty clinics or prospective volunteer brain donor studies. Participants ranged from 23 to 111 years, providing substantial representation below age 80, and more than one-third of participants with race information were Black or Brown, a population almost entirely absent from existing population-based autopsy series and from the cohorts used to derive biomarker thresholds. Recruitment occurred through a municipal death verification service rather than lifetime enrollment in a dementia research program, and prior work found that the age and sex distributions of BAS-USP participants resembled citywide deaths ^17^. Trained gerontologists obtained standardized cognitive and functional information from the most knowledgeable informant for the final 3 months of life.

The ability to distinguish CDR 0 from CDR 0.5 close to death is particularly valuable when estimating how often substantial pathology is clinically silent or accompanied by subtle impairment ^29^. Linking this assessment with comprehensive neuropathology directly characterizes amyloid, tau, neuritic plaques, and coexisting lesions in the same brain, and it provides cognitive information from age 23 onward, whereas population-scale biomarker studies have not assessed cognition below age 70 years ^13^.

### Limitations

This study has limitations. Functional cognitive status was based on a knowledgeable informant rather than direct examination or neuropsychological testing, although the standardized protocol has been validated against clinical consensus diagnosis ^29^. Requiring a knowledgeable informant with regular contact in the 6 months before death excluded socially isolated decedents, among whom cognitive impairment, alcohol use, and vascular disease are more frequent; this exclusion is likely to have biased estimates of impairment downward. Brains with large acute lesions, including extensive hemorrhages, infarctions, and tumors, were retained for determination of the cause of death and were therefore unavailable for standardized sampling. Because those are the brains most likely to carry contributing cerebrovascular pathology, the observed 20.0% frequency of contributing coexisting pathology should be read as a lower bound rather than a point estimate, and the contribution of coexisting disease to impairment is correspondingly conservative. The frequency estimates apply most directly to eligible decedents in the Sao Paulo municipal autopsy service source population. Recruitment required weekday staff availability, an informant, family consent, and suitable tissue, and estimates were not weighted to all city deaths or to living patients selected for biomarker testing. The study lacked paired antemortem plasma, cerebrospinal fluid, or PET measurements and therefore cannot directly estimate assay-specific sensitivity, specificity, or predictive values; the reported predictive values are exploratory and assume that externally derived sensitivity and specificity transfer to this population. The Thal phase and Braak stage describe anatomic distribution rather than a uniform quantitative measure of lesion burden, and biomarker signals may vary within a given phase or stage. The contributing coexisting pathology composite identified processes with sufficient burden or distribution to plausibly contribute to impairment but did not estimate each process separately.

### Conclusions

Population-based neuropathology provides age-specific tissue frequencies, reveals discordant and lower-burden amyloid and tau states, and clarifies how coexisting brain diseases complicate symptom attribution. Coexisting brain disease, undetectable by current amyloid and tau biomarkers, accompanied cognitive impairment as often as advanced AD pathology did. These data can strengthen the interpretation of positive and negative AD biomarker results as blood testing expands.

## Supporting information

Supplementary Online Content

## Data Availability

All data produced in the present study are available upon reasonable request to the authors.

## Article Information

### Funding/Support

The Biobank for Aging Studies was funded by Fundação de Amparo à Pesquisa do Estado de São Paulo (2006/55318, 2009/09134-4, 2012/07526-5, and 2024/03917-7), Conselho Nacional de Desenvolvimento Científico e Tecnológico (466763/2014-0, 465355/2014-5, and 403502/2020-9), and the Alzheimer’s Association (AARG-20-678884 and 24CBIDR-1185483). Dr Suemoto is supported by a research productivity grant from the Conselho Nacional de Desenvolvimento Científico e Tecnológico. Dr Grinberg receives support from the National Institutes of Health (K24AG053435 and R01AG075802).

### Role of the Funder/Sponsor

The funders had no role in the design and conduct of the study; collection, management, analysis, and interpretation of the data; preparation, review, or approval of the manuscript; and decision to submit the manuscript for publication.

### Data Access, Responsibility, and Analysis

Drs Grinberg and Suemoto have full access to all study data and take responsibility for its integrity and the accuracy of the analysis. Dr Pereira conducted the data analysis and is responsible for it.

### Author Contributions

Drs Pereira, Suemoto, and Grinberg had full access to all of the data in the study and take responsibility for the integrity of the data and the accuracy of the data analysis. Concept and design: Pereira, Yaffe, Suemoto, Grinberg. Acquisition, analysis, or interpretation of data: Leite, Paes, Silva, Ururahy, Moraes, Padrao, Pasqualucci, Rodrigues, Naslavsky, Ferretti-Rebustini, Nitrini, Ferrioli, Suemoto, Grinberg. Drafting of the manuscript: Drs Pereira and Grinberg. Statistical analysis: Dr Pereira. Obtained funding: Drs Suemoto, Grinberg, Leite, Nitrini, and Pasqualucci. Critical revision of the manuscript for important intellectual content: All authors. Supervision: Drs Grinberg and Suemoto.

### Data Sharing Statement

De-identified participant-level data and the data dictionary will be available to qualified researchers upon reasonable request, subject to ethics committee approval and applicable data use agreements.

### Conflict of Interest Disclosures

The authors declare no competing interests.

### Ethics

This study has been approved by the Ethics Committee of the University of Sao Paulo.

## Notes

### Competing Interest Statement

The authors have declared no competing interest.

### Author Declarations

This study has been approved by the Ethics Committee of the University of Sao Paulo.

