## Supplementary Online Content for "Amyloid and Tau Neuropathologic Stage, Coexisting Brain Disease, and Cognitive Status in a Population-Based Autopsy Cohort"

**eMethods**. Study Design, Clinical Assessment, Neuropathologic Assessment, and Detailed Statistical Analysis

**eResults**. Detailed Results

**eReferences**. List of references used only in the supplementary online content.

**eFigure 1**. Analytic Cohort Compared With All Deaths in the Municipality
**eFigure 2**. Joint Amyloid and Tau Pathology States and Clinical Dementia Rating Status Among Participants stratified by age
**eFigure 3**. Age-Specific Frequency of Combined Biomarker-Detectable Amyloid and Tau Pathology
**eFigure 4**. Prevalence of Combined Biomarker Level Amyloid and Tau Pathology and Clinical Dementia Rating Status by Age and Race Group
**eFigure 5**. Joint Amyloid and Tau Pathology States and Clinical Dementia Rating Status Among Participants Without Contributing Coexisting Pathology (n = 2170)
**eFigure 6**. Relative Importance, Sensitivity Analyses, and Validation of *m0* model (n = 1518) for Clinical Dementia Rating Sum of Boxes Scores
**eFigure 7**. Relative Importance, Sensitivity Analyses, and Validation of *m1* model (n = 2711) for Clinical Dementia Rating Sum of Boxes Scores

**eTable 1**. Demographic, Clinical, and Neuropathologic Characteristics by Braak Neurofibrillary Tangle Stage and Thal Amyloid Phase in the Full Cohort (n = 2711)
**eTable 2**. Demographic, Clinical, and Neuropathologic Characteristics by Braak Neurofibrillary Tangle Stage and CERAD Neuritic Plaque Score (n = 2711)
**eTable 3**. Demographic, Clinical, and Neuropathologic Characteristics by Braak Neurofibrillary Tangle Stage and Thal Amyloid Phase Among Female Participants (n = 1340)
**eTable 4**. Demographic, Clinical, and Neuropathologic Characteristics by Braak Neurofibrillary Tangle Stage and Thal Amyloid Phase Among Male Participants (n = 1371)
**eTable 5**. Demographic, Clinical, and Neuropathologic Characteristics by Braak Neurofibrillary Tangle Stage and Thal Amyloid Phase Among Participants Without Contributing Coexisting Pathology (n = 2170)
**eTable 6**. Coefficient Estimates from Candidate Multivariable Linear Regression Models of Clinical Dementia Rating Sum of Boxes
**eTable 7**. Exploratory Positive and Negative Predictive Values for a Plasma Phosphorylated Tau 217 Assay by Age Stratum, Using Tissue-Derived Frequency

This supplementary material has been provided by the authors to give readers additional information about their work.

### SUPPLEMENTARY MATERIALS

Amyloid and Tau Neuropathologic Stage, Coexisting Brain Disease, and Cognitive Status in a Population-Based Autopsy Cohort

#### eMethods: Study Design, Clinical Assessment, and Neuropathologic Assessment

#### Study Source, Recruitment, and Cohort Derivation

Biobank for Aging Studies at the University of Sao Paulo (BAS-USP) is an ongoing postmortem research program established in 2004 adjacent to the Sao Paulo Autopsy Service. The autopsy service is centralized for the city and receives nontraumatic deaths that require medical verification of the cause or sequence leading to death. The service performs approximately 15 000 autopsies annually. Referral to the service, therefore, occurs independently of research interest or prior willingness to donate. In the published 2004 through 2014 source population, 16.0% of all city deaths underwent autopsy at the service, and the age and sex distributions of BAS-USP participants resembled those of citywide deaths [1]. This autopsy rate is among the highest reported for any medical autopsy service worldwide. Because the published comparison covers only 2004 through 2014, we repeated it for the full study period using municipal mortality records from the Brazilian Mortality Information System (Sistema de Informacoes sobre Mortalidade). To assess the demographic representativeness of this specific study cohort relative to the citywide mortality population, we generated 10 000 bootstrap samples from the citywide mortality database, each matched to the study cohort size. Comparisons showed no evidence of differences in sex proportions after false discovery rate (FDR) adjustment ($\chi^2$ test, P = .90) or in age distributions among individuals older than 50 years when stratified by sex (females, P = .68; males, P = .78). The density of age at death to both sexes is presented in **eFigure 1**. The term population-based refers to this municipal autopsy service sampling frame rather than to a census or probability sample of all deaths in Sao Paulo. Because the brain bank enrolled a small fraction of autopsies performed at the service, representativeness is demonstrated empirically by the comparison in **eFigure 1**.

Recruitment occurred during weekday daytime periods when BAS-USP staff were available; recruitment windows comprised approximately **12** hours per week on **5** days and did change slightly over the study period when needed to accommodate external factors, like the COVID-19 epidemics in 2020 and 2021 (the autopsy service only accepted COVID-19 related deaths, and over the years, the autopsy service reorganized to perform most autopsies during the morning period. Staff first reviewed autopsy service information and study eligibility. Families were approached only after the decedent met eligibility criteria. Eligible decedents were aged 18 years or older, had a postmortem interval shorter than 24 hours, and had a legally authorized knowledgeable informant. The informant was a family member or close contact who had at least weekly contact with the decedent during the 6 months before death and could provide clinical and functional information. Written consent was obtained from the legally authorized representative before study procedures [2].

Within recruitment periods, eligibility and approach were not based on cognition, education, socioeconomic position, race, neighborhood, or referral to a clinical research program.

#### Postmortem Clinical Interview and Variable Ascertainment

After consent, a trained gerontologist conducted a standardized semi-structured interview with the most knowledgeable informant. The interview covered 3 domains: sociodemographic characteristics, medical and lifestyle history, and cognitive, functional, and neuropsychiatric status [2]. Questions about cognition and function referred to the decedent's status during the 3 months before death to reduce confounding from terminal illness and the immediate dying process.

Sociodemographic variables included age at death, sex, years of formal education, and race. Age and sex were obtained from official or autopsy service records. Education and race were reported by the informant. Race response options followed the Brazilian Institute of Geography and Statistics' categories of White, Black, Pardo, Indigenous, and Asian. Black and Pardo participants were combined as Black/Brown for the descriptive analyses, consistent with previous BAS-USP studies [2]. No participant was classified as Indigenous. These definitions are also stated in the main text Methods, as required for the reporting of race and ethnicity. Race information was available for 2061 participants; education was available for 2066. Race and education were missing when the informant did not know or declined to answer.

The medical and lifestyle portion of the interview collected information about physician-diagnosed chronic conditions, smoking, and alcohol use. These variables were not primary exposures in the current analysis but were components of the standardized interview.

The Clinical Dementia Rating, or CDR, was administered using the informant section. The 6 domains are rated for severity, and the global CDR algorithm yields a score of 0, 0.5, 1, 2, or 3 [3]. The CDR Sum of Boxes, or CDR-SB, is the sum of the 6 domain ratings and ranges from 0 to 18. For the current analysis, a global CDR score of 0 indicated no identified cognitive or functional impairment. A global CDR greater than 0, including a score of 0.5, indicated cognitive or functional impairment. **eTables 1 through 5** also report the proportion with a global CDR of 1 or higher, a dementia-level threshold, to show that the pattern across the Thal phase and Braak stage is not driven by the inclusion of very mild impairment (*i.e.*, CDR 0.5). In a validation study using the same informant protocol, the postmortem assessment had 86.6% sensitivity and 84.4% specificity for dementia and 93.7% specificity for normal cognition compared with specialist consensus in a memory clinic [4].

#### APOE Genotyping

APOE genotyping was performed when DNA was available using allele-specific real-time polymerase chain reaction for rs429358 and rs7412, with duplicate assays as previously described [2,5]. Genotypes were classified as ε2/ε2, ε2/ε3, ε2/ε4, ε3/ε3, ε3/ε4, or ε4/ε4. APOE ε4 carrier status was defined by the presence of at least 1 ε4 allele.

#### Brain Processing and Histologic Evaluation

Brain tissue was obtained within 24 hours after death. One hemisphere was fixed in 4% buffered paraformaldehyde, and selected samples from the opposite hemisphere were frozen at -80 °C. Fixed tissue was paraffin-embedded and cut at 5 µm. Systematic sampling included frontal, temporal, parietal, occipital, and cingulate cortices; hippocampal formation; amygdala; basal ganglia; thalamus; midbrain; pons; medulla; and cerebellum [2].

All sampled sections were stained with hematoxylin and eosin. Immunohistochemistry was performed on selected sections using antibodies against amyloid β (4G8; 1:10 000; BioLegend, catalog 800701), phosphorylated tau (AT8; 1:400; Invitrogen, catalog MN1020), TDP-43 (1:500; BioLegend, catalog 829901), and α-synuclein (LB509; 1:500; Sigma-Aldrich, catalog MABN824). Neuropathologic diagnoses and staging followed internationally accepted criteria and were performed by experienced neuropathologists without knowledge of the clinical interview findings [2]. All slides and tissue blocks were retained, permitting retrospective review when diagnostic criteria were updated.

#### Operationalization of Alzheimer Disease Neuropathologic Measures

The thal amyloid phase characterizes the anatomic distribution of parenchymal amyloid β plaques rather than plaque quantity. The phase assigned to each participant was the highest phase supported by the regional distribution across sampled tissue [6].

Braak neurofibrillary tangle stage characterizes the hierarchical distribution of phosphorylated tau-positive neurofibrillary tangles and neuropil threads [7]. The stage assigned to each participant was the highest stage supported across sampled regions. Braak stage reflects topographic spread and does not quantify total tangle density within a stage.

The Consortium to Establish a Registry for Alzheimer Disease, or CERAD, neuritic plaque score measures the semiquantitative neocortical density of plaques containing dystrophic neurites in selected cortical regions. Scores were classified as none, sparse, moderate, or frequent [8,9]. The unadjusted plaque density category was analyzed.

The study retained the original Thal phases and Braak stages and analyzed the CERAD neuritic plaque scores separately rather than collapsing the 3 measures into a single Alzheimer disease neuropathologic change category [9].

For prespecified summary analyses, biomarker-detectable amyloid pathology was defined as Thal phase 3 to 5, biomarker-detectable tau pathology as Braak stage IV to VI, and combined biomarker-detectable amyloid and tau pathology as meeting both thresholds. The term "biomarker-detectable" denotes stages at which current amyloid and tau assays are expected to yield positive results. The correspondence is inferred from published autopsy-validation studies and was not measured in this cohort, since no participant had antemortem biomarker data. These were study-specific clinicopathologic categories rather than universal positivity thresholds for an individual imaging or fluid biomarker. Lower-burden and discordant states remained visible in the full Thal phase by Braak stage matrix, and because detection thresholds will change as assays improve.

#### Operationalization of Contributing Coexisting Pathology

Contributing coexisting pathology was a study-specific composite intended to identify non-Alzheimer disease processes with sufficient burden or distribution to contribute materially to cognitive or functional impairment. A participant was classified as having a contributing coexisting pathology when at least 1 of the following conditions was present.

> Contributing cerebrovascular pathology was defined using the BAS-USP operational criteria for vascular dementia, informed by the Newcastle categorization. Vascular dementia was assigned when at least 1 of the following was present: an extensive chronic infarct larger than 1 cm; a lacunar infarct smaller than 1 cm in a strategic region, including the thalamus, front cingulate cortex, basal forebrain, caudate nucleus, mesial temporal region, or angular gyrus; or lacunar infarcts involving 3 or more cortical regions. Small-vessel disease alone was insufficient for this diagnosis. Cerebral amyloid angiopathy was evaluated separately and was not included in the contributing coexisting pathology composite [2,10].
>
> Limbic-predominant age-related TDP-43 encephalopathy neuropathologic change was assessed by TDP-43 immunohistochemistry. Stage 1 was defined as pathology limited to the amygdala, stage 2 as pathology in the amygdala and hippocampus, and stage 3 as additional involvement of the middle frontal gyrus. Only stages 2 and 3 were included in the contributing coexisting pathology composite [11].
>
> Lewy-type pathology was assessed by α-synuclein immunohistochemistry and staged using Braak Parkinson disease criteria [12]. Only stages 5 and 6 were included in the contributing composite because these stages represent neocortical spread; stages 1 through 4 were not classified as contributing for this analysis [13].
>
> Hippocampal sclerosis was defined as disproportionate severe pyramidal neuronal loss and gliosis in the CA1 region and subiculum and was scored as present or absent [2,14]. Primary frontotemporal lobar degeneration associated with tau or TDP-43 pathology was included when diagnostic neuropathologic criteria were met [15].

Argyrophilic grain disease, aging-related tau astrogliopathy, cerebral amyloid angiopathy, limbic-predominant age-related TDP-43 encephalopathy neuropathologic change stage 1, Lewy body pathology below Braak Parkinson disease stage 5, and incidental vascular lesions were not included in the primary composite. Primary age-related tauopathy was represented within the Thal phase by Braak stage matrix rather than classified as a separate coexisting pathology.

Analyses were conducted in the full cohort and repeated after excluding all participants who met the definition of contributing coexisting pathology. Because each component threshold was set conservatively and because brains with large acute lesions were unavailable for standardized sampling, the resulting frequency of contributing coexisting pathology (20.0%) is likely a lower bound.

### eMethods: Detailed Statistical Analysis

#### Analysis Framework and Estimands

Analyses were organized around the 3 biomarker interpretation questions specified in the Introduction. To inform target detection, we estimated the overall and age- and sex-specific frequency of biomarker-detectable amyloid pathology, biomarker-detectable tau pathology, and combined biomarker-detectable amyloid and tau pathology. To assess pathologic characterization, we described the full Thal phase by Braak stage matrix, summarized CERAD neuritic plaque burden separately, and classified participants into 4 mutually exclusive states: neither threshold, amyloid only, tau only, and both thresholds. To assess clinical attribution, we estimated the proportion with CDR greater than 0 within matrix cells and summary states, cross-classified combined biomarker-detectable amyloid and tau pathology with contributing coexisting pathology and repeated the analyses after excluding participants with contributing coexisting pathology.

The primary descriptive representation was the complete Thal phase by Braak stage matrix. The prespecified summary estimand was the frequency of combined biomarker-detectable amyloid and tau pathology at death in the municipal autopsy service source population. These estimates were intended as neuropathologic priors and do not directly estimate the sensitivity, specificity, positive predictive value, or negative predictive value of a positron emission tomography (PET), cerebrospinal fluid, or plasma assay.

#### Analysis Populations, Missing Data, and Sample Size

The primary analytic cohort included 2711 participants with complete assessments of Thal phase, Braak stage, CERAD neuritic plaque score, global CDR, CDR-SB, and contributing coexisting pathology. Descriptive analyses used all participants with data available for the variables included in each analysis. Race was available for 2061 participants, education for 2066, and APOE genotyping for 1518. Missing values were not imputed. No a priori sample size calculation was performed. The analytic cohort included all available eligible participants with complete primary clinicopathologic variables during the study period.

Multivariable linear regression models were used to evaluate independent associations with CDR-SB, analyzed as a continuous outcome ranging from 0 to 18. Multivariable models used complete observations for all variables included in the corresponding model. Age at death was modeled in decades, and education was modeled in years. Sex was entered as female versus male, APOE ε4 status as carrier versus noncarrier, and contributing coexisting pathology as present versus absent. No interaction terms were included in the primary model.

The fully adjusted primary model (*m0*) required complete education and APOE data and therefore included **1518** participants, or **56**% of the analytic cohort. Because the variables included differed across candidate models, their analytic samples could also differ. A second model (*m1;* n = **2711**, **100**% of the cohort), excluding education and APOE ε4 status due to the missing data and lower or null participation in *m0*, was included.

#### Descriptive Statistics and Table Comparisons

Cohort characteristics were summarized as mean (SD) for continuous variables and No. (%) for categorical variables. Unadjusted comparisons by global CDR status used 2-sample t tests for continuous variables, and Pearson $\chi^2$ tests for categorical variables. Fisher exact tests were used for sparse contingency tables when appropriate. These comparisons were descriptive and were not used to select covariates for the multivariable models.

P values in Table 1 and eTables 1 through 5 were descriptive, 2-sided, and unadjusted. The P values in **eTables 1 through 5** represent overall comparisons across Braak stages within the corresponding Thal phase or CERAD neuritic plaque score category. No adjustment was made for multiple comparisons.

#### Operational Definitions for Summary Analyses

The complete Thal phase by Braak stage matrix was retained as the primary descriptive representation, and the CERAD neuritic plaque score was analyzed separately. Biomarker-detectable amyloid pathology was defined as Thal phase 3 to 5, biomarker-detectable tau pathology as Braak stage IV to VI, and combined biomarker-detectable amyloid and tau pathology as meeting both thresholds.

The Thal phase 3 was selected as a pragmatic study threshold representing more advanced amyloid β plaque distribution that is generally associated with greater amyloid PET signal, while recognizing that amyloid PET positivity can occur at lower Thal phases depending on the tracer and analytic threshold [16,17]. Braak stage IV was selected as a pragmatic threshold because quantitative regional flortaucipir approaches can detect intermediate tau burden around this stage [18,19]. Standard clinical flortaucipir visual interpretation is most strongly associated with B3 pathology, corresponding to Braak stages V to VI [20], and autopsy studies show lower sensitivity for earlier Braak stages [21]. These categories summarize neuropathologic tissue states rather than universal positivity thresholds for individual PET, cerebrospinal fluid, or plasma assays [22,23].

Counts for the 4 mutually exclusive amyloid and tau states were generated directly from participant-level threshold indicators. Conditional concordance was summarized as the proportion meeting the tau threshold among participants meeting the amyloid threshold and the proportion meeting the amyloid threshold among participants meeting the tau threshold. No inferential tests were performed across individual matrix cells.

#### Age, Sex, and Race Group Analyses

Age at death was analyzed in descriptive strata of 23 to 50 years, 51 to 60 years, 61 to 70 years, 71 to 80 years, 81 to 90 years, and 91 to 111 years. The observed age range was 23 to 111 years. Sex was categorized as female or male. Frequency estimates were calculated within each age- and sex-stratum in the full cohort and after excluding participants with contributing coexisting pathology.

For the exploratory race group analysis, participants were classified as White or Black/Brown. The same 3 comparisons were made in the full cohort and after excluding participants with contributing coexisting pathology.

#### Relative Importance and Sensitivity Analyses

Term-level partial *R2* quantified the unique contribution of each predictor after adjustment for the remaining variables. Unique *ΔR2* quantified the reduction in total model *R2* after removing each predictor. The Lindeman, Merenda, and Gold method allocated the total explained variance among predictors by averaging their incremental contributions over all possible predictor orderings.

Heteroskedasticity-consistent HC3 covariance estimates were used for term-level inference. Participant-level bootstrap resampling was used to estimate 95% CIs and the stability of variable importance rankings.

Because CDR-SB is bounded between 0 and 18 and linear model diagnostics indicated heteroskedasticity, nonnormal residuals, and boundary-related misspecification, a quasi-binomial fractional logit model was fitted to CDR-SB divided by 18. HC3 robust term tests were used for this model. Adjusted predictions and contrasts were transformed back to the original CDR-SB scale.

Ten-fold cross-validation compared the linear and fractional logit models using mean absolute error, root mean square error, predictive *R2*, the percentage of predictions outside the valid 0 to 18 range, and calibration between mean predicted and observed CDR-SB values.

#### Prevalence exploratory analysis

In an exploratory analysis, we estimated age-stratum-specific positive and negative predictive values for a plasma p-tau217 assay by combining the observed tissue frequency of combined biomarker-detectable amyloid and tau pathology with externally derived sensitivity and specificity ranges from published autopsy-validated studies. For each age stratum and each combination of sensitivity, specificity, and frequency, predictive values were computed by the Bayes formulas PPV = (Se x Prev) / (Se x Prev + (1 - Sp) x (1 - Prev)) and NPV = (Sp x (1 - Prev)) / ((1 - Se) x Prev + Sp x (1 - Prev)) [24]. We summarized the resulting distributions with the median and the 2.5th to 97.5th percentiles. Because external sensitivity and specificity typically attenuate on transfer to new populations, we repeated the calculation after shrinking both toward 0.5 by 10%. Results are given in eTable 7 and Figure 4. Two assumptions are required and stated as limitations: that externally derived assay performance applies to this population, and that the frequency at death approximates the pretest probability in the clinical population being tested. Unlike published predictive-value estimates anchored to biomarker-defined frequency, these estimates do not use the same measurement to define both the prior and the test.

#### Software

All analyses were performed using R version 4.6.1 with packages *lmtest* for statistical models, *sandwich* for HC3, *relaimpo* for LMG and *boot* for bootstrap.

### eResults

#### Analysis Population and Data Availability

Of 2716 available participants, 3 were excluded because global CDR or CDR-SB data were unavailable and 2 because Thal phase or Braak stage data were unavailable. The final analytic cohort included 2711 participants. The mean [SD] age at death was 73.4 [14.40] years, with an observed range of 23 to 111 years. Of the 2711 participants, 1340 (49.4%) were female, 1787 (65.9%) had CDR 0, and 924 (34.1%) had CDR greater than 0 (**Figure 1 and Table 1**).

Race was available for 2061 participants, education for 2066, and APOE genotyping for 1518. Mean [SD] years of education were 5.4 [4.37], 5.9 [4.47] among participants with CDR 0, and 4.3 [3.90] among participants with CDR greater than 0. Among participants with genotyping data, 450 of 1518 (29.6%) were APOE ε4 carriers. A contributing coexisting pathology was present in 541 participants (20.0%) (**Table 1**).

#### Thal, Braak, and CERAD Distributions

The full cohort distribution of Thal amyloid phase by Braak neurofibrillary tangle stage is presented in **Figure 2A** and **eTable 1**. The Thal phase was 0 in 1318 participants (48.6%), 1 in 366 (13.5%), 2 in 226 (8.3%), 3 in 342 (12.6%), 4 in 364 (13.4%), and 5 in 95 (3.5%). Braak stage was 0 in 541 participants (20.0%), I in 403 (14.9%), II in 426 (15.7%), III in 603 (22.2%), IV in 371 (13.7%), V in 188 (6.9%), and VI in 179 (6.6%).

The distribution of CERAD neuritic plaque score by Braak stage is presented in **eTable 2**. The CERAD score was none in 1661 participants (61.3%), sparse in 380 (14.0%), moderate in 393 (14.5%), and frequent in 277 (10.2%).

Sex-specific distributions are presented in **eTables 3 and 4**. The corresponding Thal phase by Braak stage distribution among 2170 participants without contributing coexisting pathology is presented in **eTable 5** and **eFigure 5**.

#### Joint Amyloid and Tau States

Biomarker-detectable amyloid pathology, defined as Thal phase 3 to 5, was present in 801 of 2711 participants (29.5%). Biomarker-detectable tau pathology, defined as Braak stage IV to VI, was present in 738 participants (27.2%). A total of 531 participants (19.6%) met both thresholds (**Figure 2B**).

Among the 4 mutually exclusive amyloid and tau states, 1703 participants (62.8%) met neither threshold, 270 (10.0%) met the amyloid threshold only, 207 (7.6%) met the tau threshold only, and 531 (19.6%) met both thresholds (**Figure 2B**). Among participants meeting the amyloid threshold, 531 of 801 (66.3%) also met the tau threshold. Among participants meeting the tau threshold, 531 of 738 (72.0%) also met the amyloid threshold. Discordant states, in which only 1 of the 2 thresholds was met, accounted for 477 of 2711 participants (17.6%).

Figure 3B displays the combined biomarker-detectable amyloid and tau pathology group relative to all other amyloid and tau states.

#### Clinical Expression and Contributing Coexisting Pathology

The proportion of participants with CDR greater than 0 increased monotonically across Braak stage, collapsing across Thal phase: 88 of 541 (16.3%) at stage 0, 74 of 403 (18.4%) at stage I, 112 of 426 (26.3%) at stage II, 176 of 603 (29.2%) at stage III, 181 of 371 (48.8%) at stage IV, 138 of 188 (73.4%) at stage V, and 155 of 179 (86.6%) at stage VI (**eTable 1**). In the full cohort, CDR greater than 0 was present in 381 of 531 participants (71.8%) meeting both amyloid and tau thresholds and in 543 of 2180 (24.9%) in the other 3 states. The absolute difference was 46.8 percentage points (95% CI, 42.5 to 50.9 percentage points; $P < .001$).

Among participants without contributing coexisting pathology (**eTable 5 and eFigure 5**), CDR greater than 0 was present in 216 of 339 participants (63.7%) meeting both thresholds and in 339 of 1831 (18.5%) in the other 3 states. The absolute difference was 45.2 percentage points (95% CI, 39.6 to 50.4 percentage points; $P < .001$).

When combined biomarker-detectable amyloid and tau pathology and contributing coexisting pathology were cross-classified, CDR greater than 0 was present in 165 of 192 participants (85.9%) with both conditions, 216 of 339 (63.7%) with combined biomarker-detectable amyloid and tau pathology alone, 204 of 349 (58.5%) with contributing coexisting pathology alone, and 339 of 1831 (18.5%) with neither condition. Contributing coexisting pathology alone was therefore accompanied by impairment nearly as often as combined amyloid and tau pathology alone (**Figure 2C**).

Among participants meeting both amyloid and tau thresholds, the frequency of CDR greater than 0 was 85.9% among those with contributing coexisting pathology and 63.7% among those without contributing coexisting pathology. The absolute difference was 22.2 percentage points (95% CI, 14.7 to 28.9 percentage points; P < .001).

#### Age- and Sex-Specific Estimates

The frequency of combined biomarker-detectable amyloid and tau pathology increased across age groups (**eFigures 2 and 3**). Frequency was 0% among participants aged 23 to 50 years, 2.0% at ages 51 to 60 years, 5.5% at ages 61 to 70 years, 14.5% at ages 71 to 80 years, 36.5% at ages 81 to 90 years, and 45.9% among participants aged 91 to 111 years.

Frequency estimates did not differ significantly between female and male participants at ages 70 years or younger and were higher among female participants at older ages. Among participants aged 91 to 111 years, frequency was 54.2% in female participants and 26.5% in male participants. The age pattern was similar after excluding participants with contributing coexisting pathology. Across the whole cohort, the crude frequency was 376 of 1340 (28.1%) in female and 155 of 1371 (11.3%) in male participants; because female participants died at older ages, this crude difference largely reflects age, and the age-stratified estimates are the interpretable comparison.

Age- and sex-specific estimates and descriptive P values are shown in **eTables 3 and 4**.

#### Exploratory Race Group Analysis

Race information was available for 2061 participants, including 1272 (61.7%) classified as White, 753 (36.5%) as Black/Brown, and 36 (1.7%) as Asian; no participant was classified as Indigenous.

For the exploratory analysis shown in **eFigure 4**, participants were classified as White or Black/Brown. In the full cohort, combined biomarker level amyloid and tau pathology was present in approximately 20% of White participants and 13% of Black/Brown participants. After excluding participants with contributing coexisting pathology, the corresponding estimates were approximately 17% and 10%. In the full cohort, and after excluding participants with contributing coexisting pathology, the proportion of participants with combined biomarker-detectable amyloid and tau differed between White and Black/Brown, with a lower proportion in the latter (both adjusted P-values < .001). This difference was not observed when the analysis was stratified by age at death. The proportion of CDR > 0 for combined or non-combined biomarker levels did not differ statistically across groups or by age.

In the AHEAD 3-45 screening cohort, plasma phosphorylated tau 217 percentage (%p-tau217) differed across racial and ethnic groups, although its association with amyloid PET showed no significant pairwise differences across racial groups [25]. Our autopsy cohort provides complementary evidence by directly characterizing amyloid, tau, and coexisting pathologies in a racially diverse Brazilian population.

#### Multivariable Associations with CDR Sum of Boxes

In the adjusted complete-case model *m0* (n = 1518), education was inversely associated with CDR-SB but had a small contribution (β = -0.09, 95% CI -0.149 to -0.037, $P < .001$), whereas APOE ε4 carrier status was not independently associated ($P = .26$). Given their substantial missingness and limited contribution, both variables were omitted from the secondary adjusted model m1, which included all 2711 participants. In m1, Braak stage VI was associated with a 6.13-point higher CDR-SB score (95% CI, 4.932 to 7.324, $P < .001$), contributing coexisting pathology with a 4.03-point (95% CI, 3.538 to 4.517, $P < .001$), Braak stage V with 3.34-point (95% CI, 2.212 to 4.466, $P < .001$), CERAD 3 with 2.38-point (95% CI, 1.212 to 3.558, $P < .001$), and Thal phase 5 with 2.19-point (95% CI, 0.735 to 3.642, $P = .003$).

Also, Braak Stage IV (β = 1.22, 95% CI 0.407 to 2.039, $P = .003$), CERAD 2 (β = 1.48, 95% CI 0.496 to 2.459, $P = .003$), female sex (β = 0.79, 95% CI 0.404 to 1.183, $P < .001$), and 10-year increase in age at death (β = 0.06, 95% CI 0.045 to 0.079, $P < .001$) were positively associate.

Braak stages I, II, and III, CERAD 1, and Thal phases 1, 2, 3, and 4 were not statistically significantly associated or were minimally negatively associated. Complete coefficient estimates, units, reference categories, 95% CIs, P values, and analytic sample sizes for the models *m0* and *m1* are reported in **eTable 6**.

#### Relative Importance and Sensitivity Analyses

Term-level partial *R2* quantified the unique contribution of each predictor after adjustment for the remaining variables, and the Lindeman, Merenda, and Gold (LMG) method allocated the total explained variance among the *m0* and *m1* predictors. Year of education and APOE ε4 carrier status had the lowest bootstrap partial *R2* on *m0* (**Figure 3A** and **eFigure 6**). On *m1,* contributing coexisting pathology had the largest unique contribution to CDR-SB variation. Its partial R2 was 0.089 (95% bootstrap CI, 0.064 to 0.114), and its unique *ΔR2* was 0.060. Contributing coexisting pathology ranked first by partial *R2* in 95.4% of bootstrap samples. Braak stage had the second largest unique contribution, with a partial *R2* of 0.057 (95% CI, 0.038 to 0.083) and a unique *ΔR2* of 0.037. Braak stage had the largest LMG contribution to total *R2*, at 0.115, and ranked first by this measure in 97.5% of bootstrap samples. These findings indicate that Braak stage captured both unique explanatory information and information shared with other neuropathologic measures (**Figure 3B** and **eFigure 7**).

The HC3 linear and fractional logit analyses produced similar term-level conclusions. In the fractional logit model, contributing coexisting pathology was associated with a 4.03-point higher adjusted CDR-SB score compared with its absence (95% bootstrap CI, 3.41 to 4.65). Compared with Braak stage 0, stages IV, V, and VI were associated with adjusted differences of 1.22, 3.34, and 6.13 CDR-SB points, respectively. Category-specific Thal phase estimates were nonmonotonic despite evidence from the global categorical term test (**eFigure 7**).

#### Model Validation

The fractional logit model *m1* produced no predictions below 0 or above 18. The linear model *m1* produced predictions above 18 in 0.2% of cases, and 9.1% of its cross-validated predictions were below 0 (**eFigure 7E**). Calibration across predicted risk groups was similar for the 2 models, with the fractional logit model maintaining all predictions within the valid CDR-SB range (**eFigure 7F**).

#### Positive Predictive Value

This analysis is exploratory and requires 2 assumptions. First, that externally derived sensitivity and specificity transfer to this population. Second, that frequency at death approximates the pretest probability in the clinical population being tested; frequency at death in an autopsy source population is not equivalent to frequency among living patients presenting for biomarker evaluation. Unlike published predictive-value estimates in which the frequency used as the prior was itself derived from the biomarker under evaluation, the prior used here is derived from neuropathologic examination and is therefore independent of the assay.

Combining the observed tissue frequencies with externally [24] derived reported sensitivity of 94% and specificity of 94% ranges using moderate-to-frequent neuritic plaques for plasma p-tau217, the estimated positive predictive value rose with age from 47.7% at ages 61 to 70 years to 72.7% at ages 71 to 80 years and 93.0% at ages 91 to 111 years, while the negative predictive value fell from 99.6% to 94.9%. The positive predictive values dropped to 21.4%, 44.1%, and 78.9% for the same age strata when a sensitivity of 94% and specificity of 79% were used, considering the broader intermediate-to-high neuritic plaques category from the same report [24]. At this operating point, the positive predictive value reached 50% only once the frequency of the target pathology exceeded 6.0% for sensitivity/specificity 94%/94%, and 18.3% for 94%/79%. Values for all age strata, including optimism-corrected estimates and the biomarker-anchored comparison, are given in **Figure 4** and **eTable 7**. Because the prior is tissue-derived, these estimates are not subject to the circularity that arises when frequency is estimated from the same biomarker being evaluated.

**eFigure 1. Analytic Cohort Compared With All Deaths in the Municipality.**

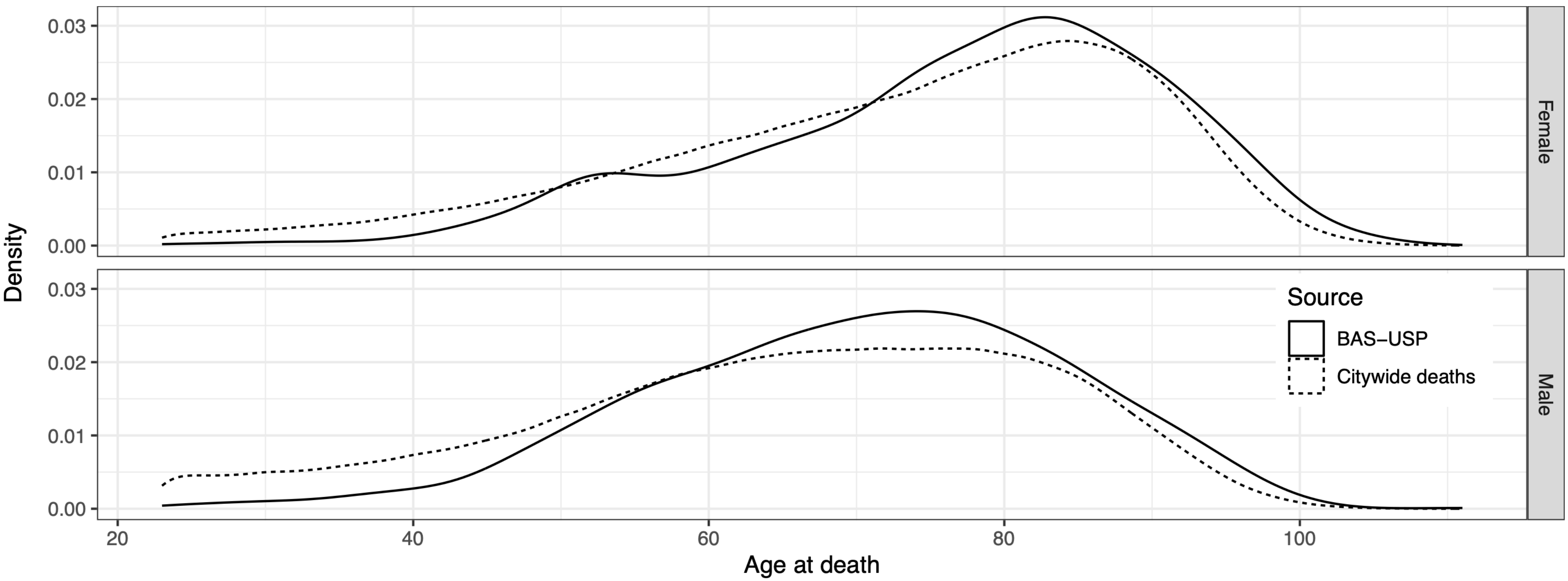


Distribution of age at death and sex in the analytic cohort (n = 2711) and in all deaths in the municipality. Municipal mortality data were obtained from the Brazilian Mortality Information System.

**eFigure 2. Joint Amyloid and Tau Pathology States and Clinical Dementia Rating Status Among Participants stratified by age.**

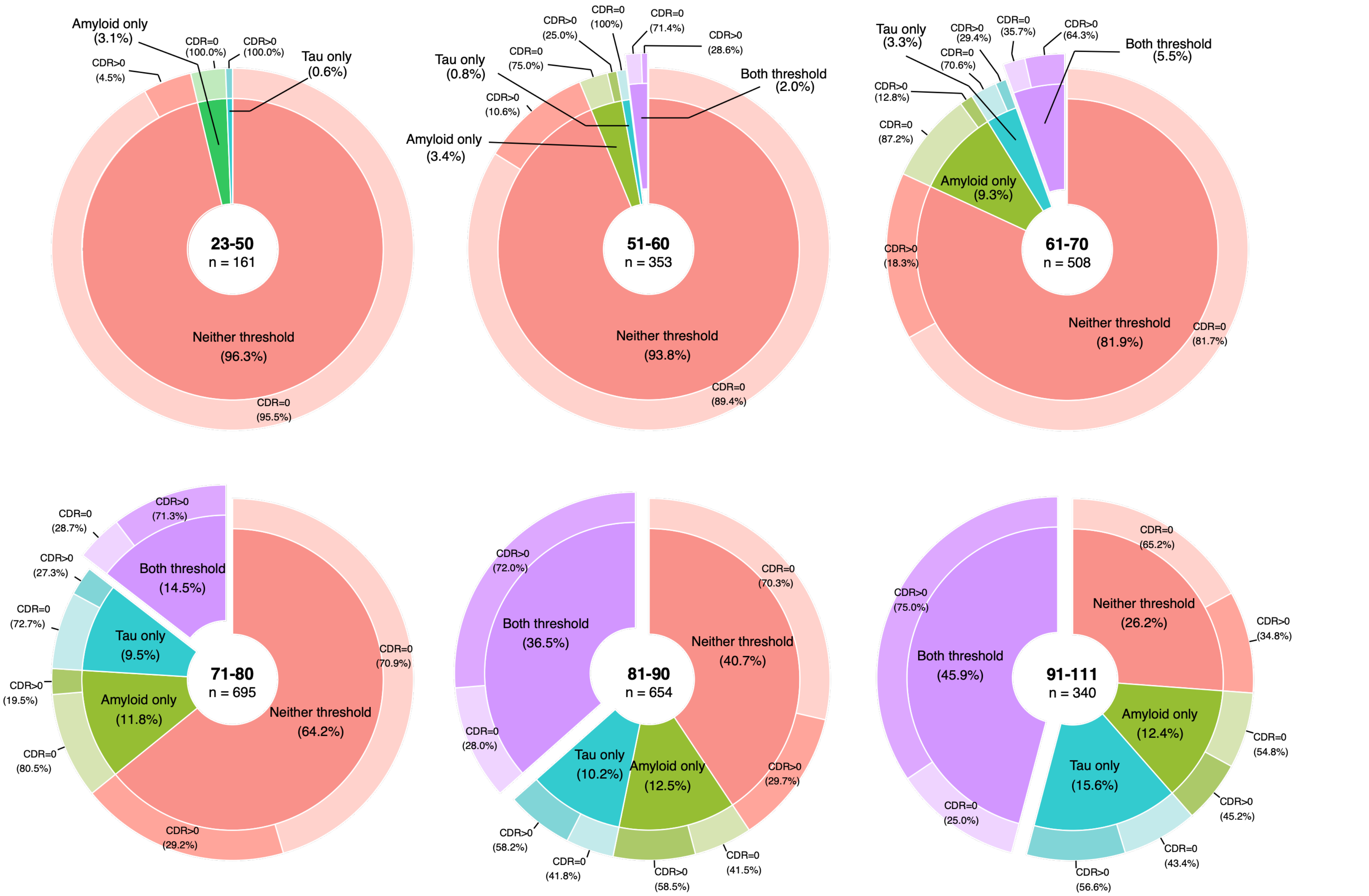


Joint distribution of Thal amyloid phase and Braak neurofibrillary tangle stage among participants stratified by age. Each pie chart displays the number of participants, the percentage of the age group, and the percentage with CDR greater than 0 with the neither threshold (pink-ish), biomarker-detectable amyloid pathology (green-ish), biomarker-detectable tau pathology (turquois blue-ish), and both biomarker-detectable threshold (purple-ish). Bright color in outer circles indicates CDR 0, and darker color indicates CDR greater than 0.
Abbreviations: CDR, Clinical Dementia Rating.

**eFigure 3. Age-Specific Frequency of Combined Biomarker-Detectable Amyloid and Tau Pathology.**

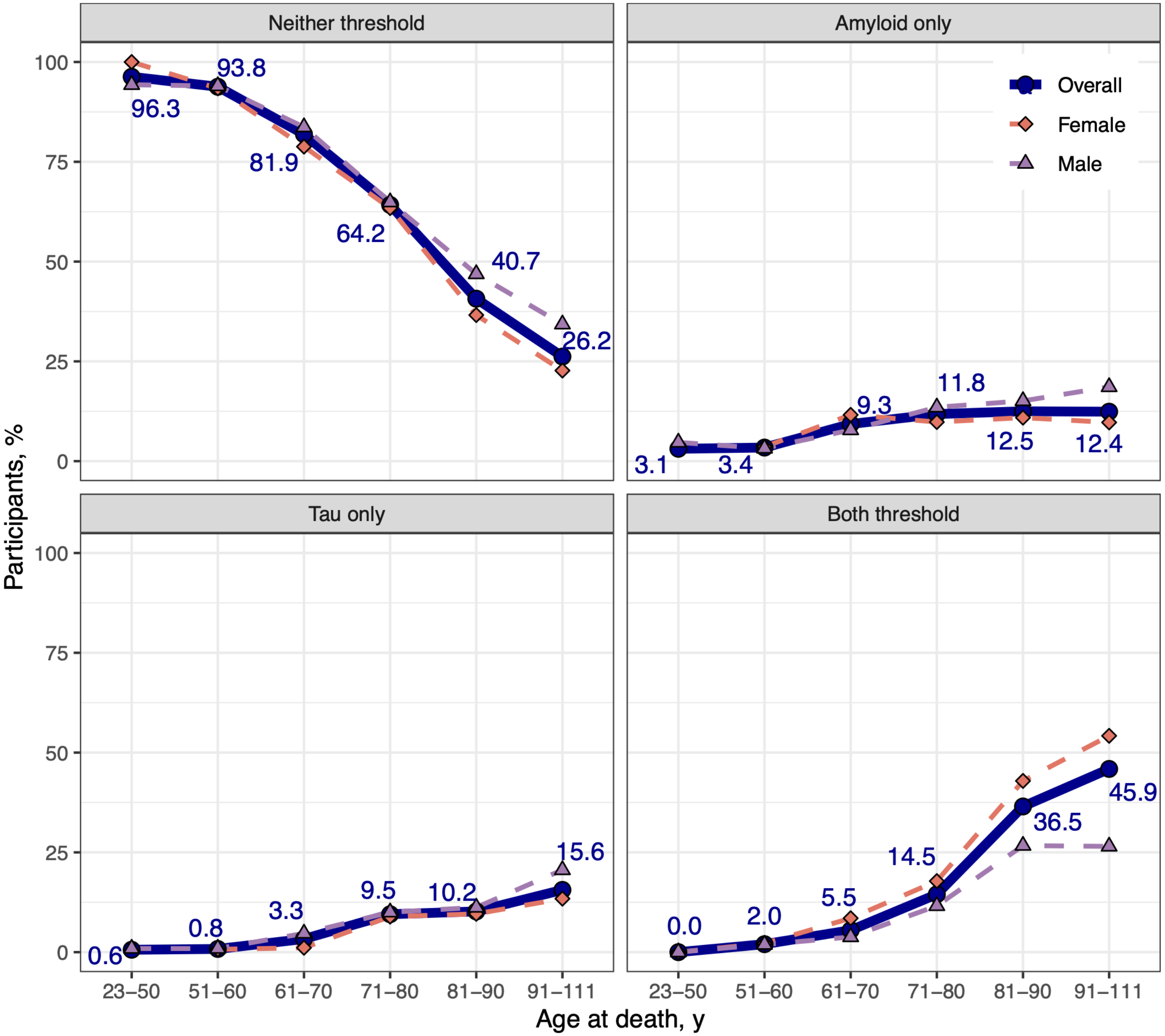


Frequency of the cases with neither (top-left), only-amyloid (top-right), only-tau (bottom-left) and both threshold (bottom-right) biomarker-detectable distribution pathology, by age at death, overall (dark blue solid line and circles) and by sex (female in red-ish dashed line and diamonds; and male in purple-ish dashed line and triangles). The threshold is defined as the Thal phase 3 to 5 and the Braak stage IV to VI.

#### eFigure 4. Prevalence of Combined Biomarker Level Amyloid and Tau Pathology and Clinical Dementia Rating Status by Age and Race Group

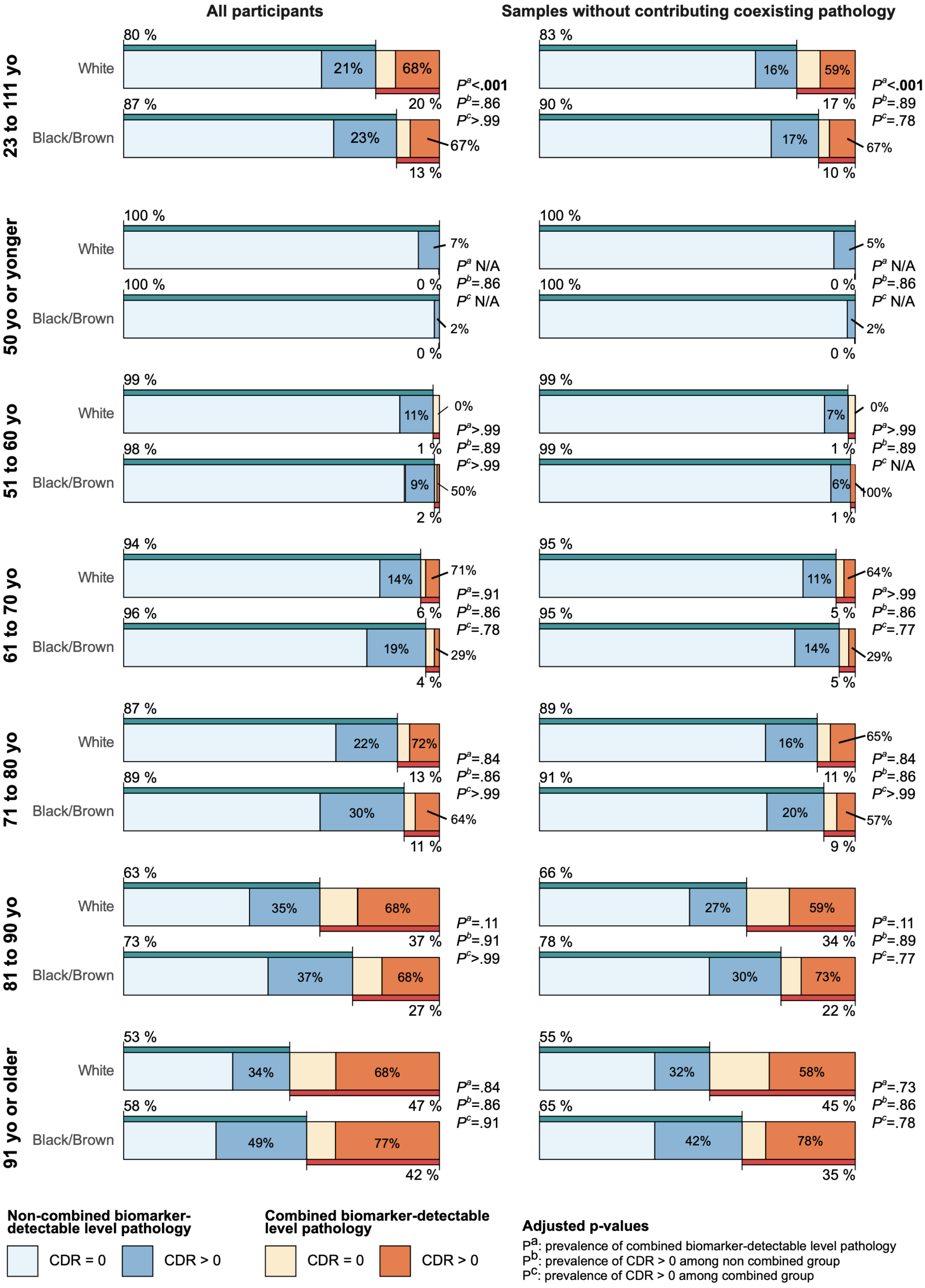

**eFigure 5. Joint Amyloid and Tau Pathology States and Clinical Dementia Rating Status Among Participants Without Contributing Coexisting Pathology (n = 2170)**

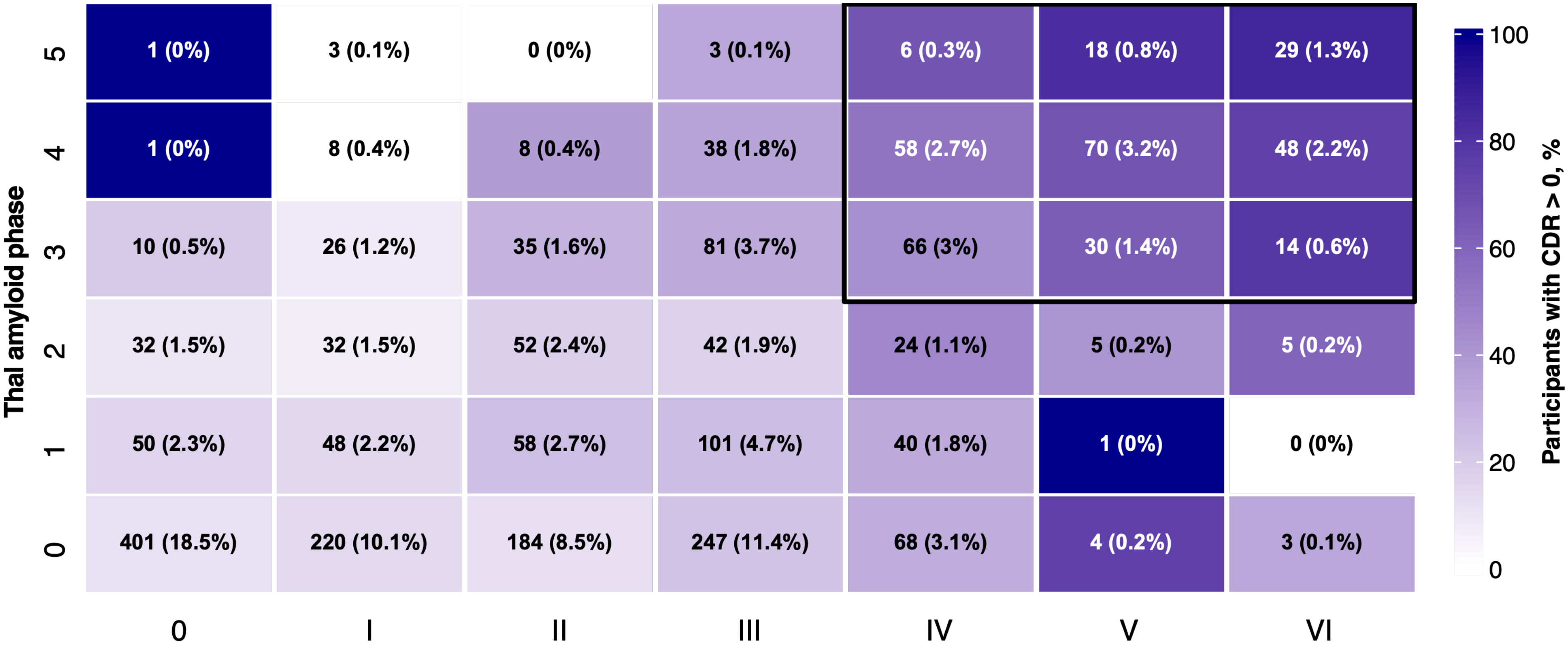


Joint distribution of Thal amyloid phase and Braak neurofibrillary tangle stage among 2170 participants without contributing coexisting pathology. Each cell displays the number of participants, the subgroup percentage and the percentage with CDR greater than 0. Background shading indicates the percentage with CDR greater than 0.

#### eFigure 6. Relative Importance, Sensitivity Analyses, and Validation of *m0* model (n = 1518) for Clinical Dementia Rating Sum of Boxes Scores

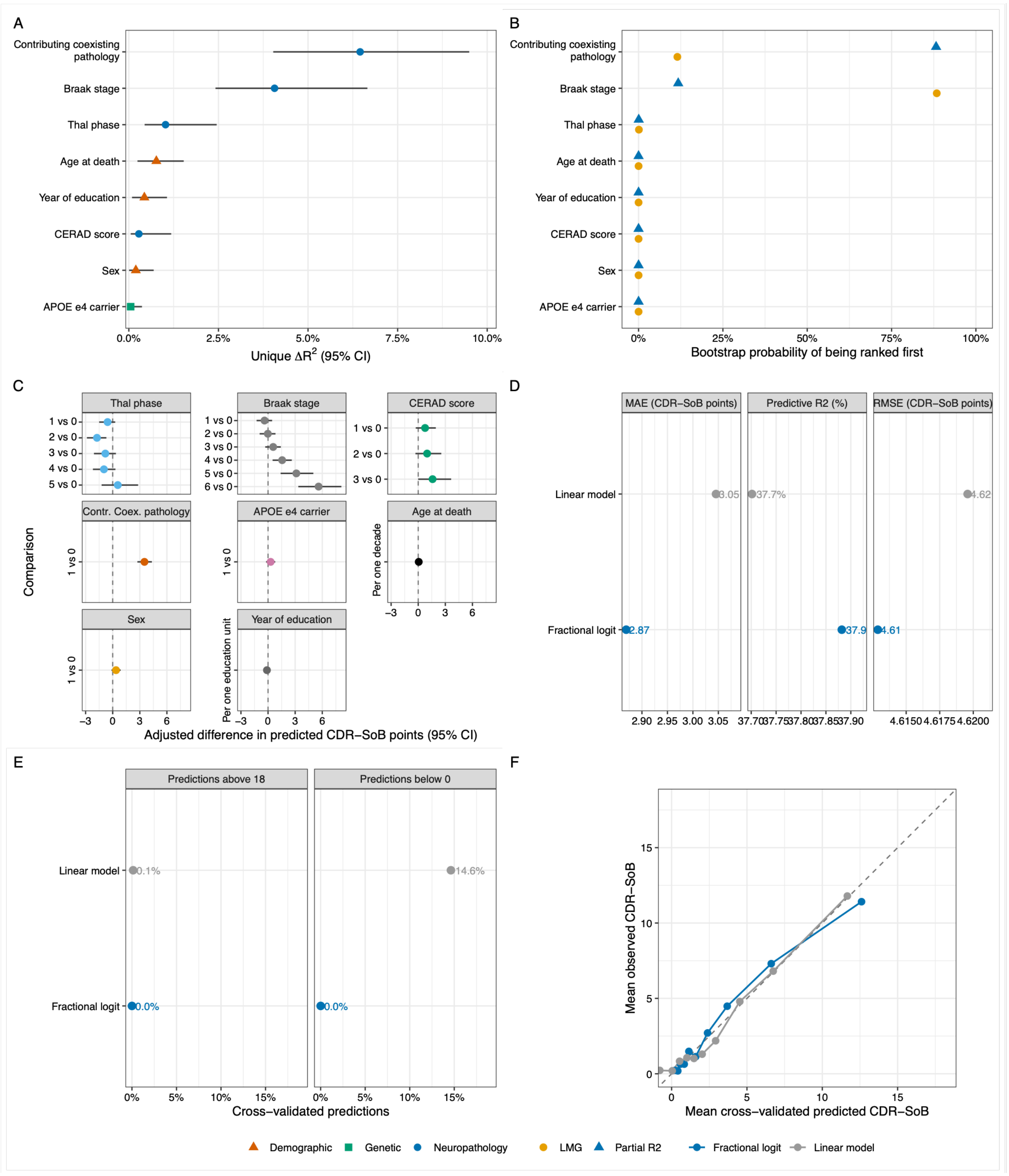


A, Unique reduction in total R² after removal of each predictor, with percentile bootstrap 95% CIs. B, Bootstrap probability that each predictor ranked first by partial R² or by Lindeman, Merenda, and Gold contribution. C, adjusted differences in predicted CDR-SB scores from the fractional logit model, transformed back to the original 0 to 18 scale. The Thal phase, Braak stage, and CERAD neuritic plaque score categories are compared with category 0. Contributing coexisting pathology is compared with its absence, APOE ε4 carrier status with noncarrier status, and female sex with male sex. Age at death is modeled per 10-year increase, and education per 1-year increase. D, Ten-fold cross-validated mean absolute error, predictive R², and root mean square error for the linear and fractional logit models. E, Percentage of cross-validated predictions above 18 or below 0. F, Decile binned cross-validated calibration. The dashed line indicates perfect agreement between the mean predicted and observed scores.

Abbreviations: APOE, apolipoprotein E; CDR-SB, Clinical Dementia Rating Sum of Boxes; CERAD, Consortium to Establish a Registry for Alzheimer Disease; CI, confidence interval; LMG, Lindeman, Merenda, and Gold; MAE, mean absolute error; RMSE, root mean square error; R², coefficient of determination.

**eFigure 7. Relative Importance, Sensitivity Analyses, and Validation of *m1* model (n = 2711) for Clinical Dementia Rating Sum of Boxes Scores**

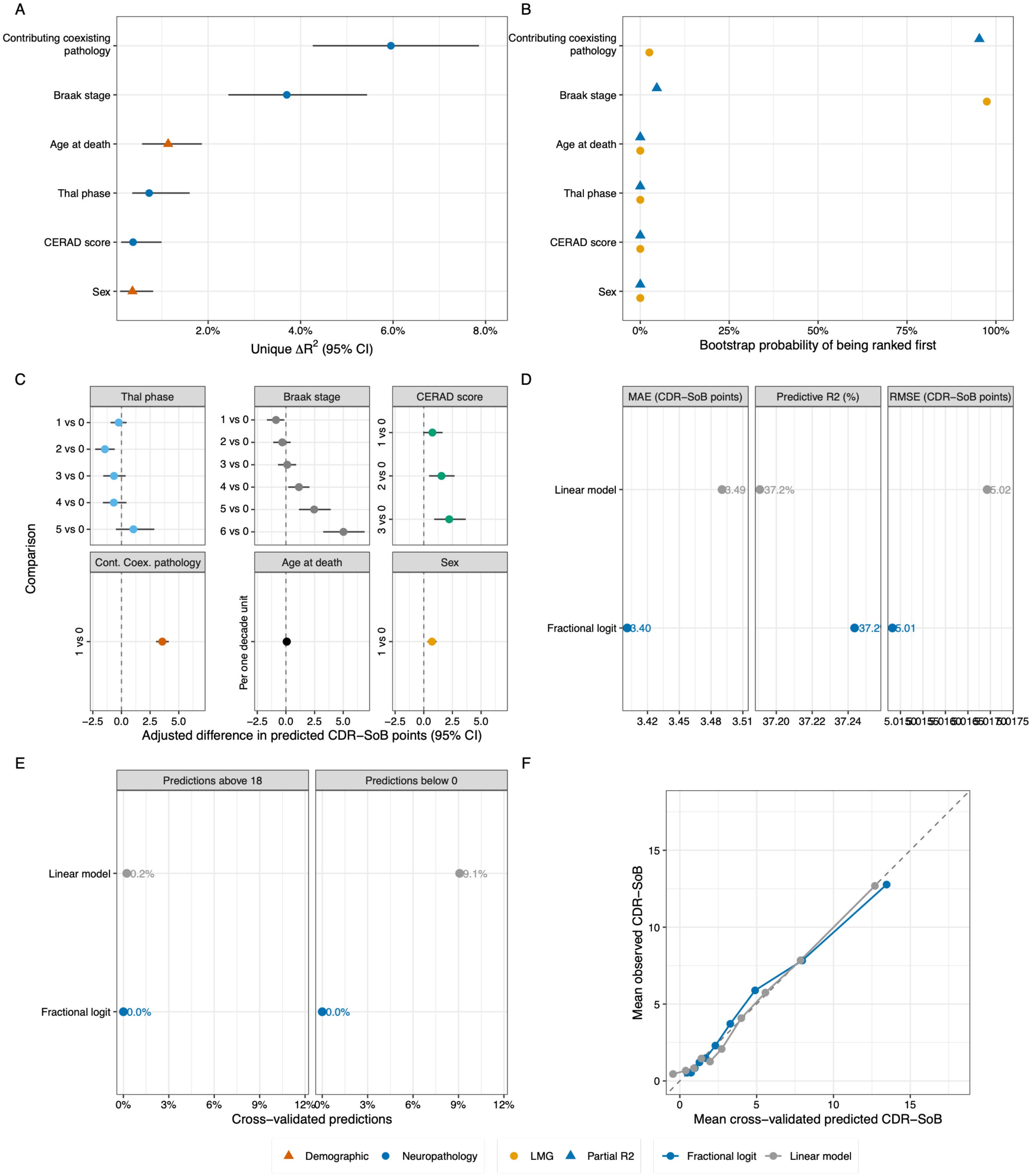


A, Unique reduction in total R² after removal of each predictor, with percentile bootstrap 95% CIs. B, Bootstrap probability that each predictor ranked first by partial R² or by Lindeman, Merenda, and Gold contribution. C, adjusted differences in predicted CDR-SB scores from the fractional logit model, transformed back to the original 0 to 18 scale. The Thal phase, Braak stage, and CERAD neuritic plaque score categories are compared with category 0. Contributing coexisting pathology is compared with its absence, and female sex with male sex. Age at death is modeled per 10-year increase. D, Ten-fold cross-validated mean absolute error, predictive R², and root mean square error for the linear and fractional logit models. E, Percentage of cross-validated predictions above 18 or below 0. F, Decile binned cross-validated calibration. The dashed line indicates perfect agreement between the mean predicted and observed scores.

Abbreviations: APOE, apolipoprotein E; CDR-SB, Clinical Dementia Rating Sum of Boxes; CERAD, Consortium to Establish a Registry for Alzheimer Disease; CI, confidence interval; LMG, Lindeman, Merenda, and Gold; MAE, mean absolute error; RMSE, root mean square error; R², coefficient of determination.

#### eTable 1. Demographic, Clinical, and Neuropathologic Characteristics by Braak Neurofibrillary Tangle Stage and Thal Amyloid Phase in the Full Cohort (n = 2711)

| Variable | Total | Braak stage | | | | | | | P value |
|---|---|---|---|---|---|---|---|---|---|
| | | 0 | I | II | III | IV | V | VI | |
| **Thal phase, No. (%)** | | | | | | | | | |
| Thal 5 | 95 (3.5) | 1 (1.1) | 4 (4.2) | 0 (0) | 4 (4.2) | 9 (9.5) | 24 (25.3) | 53 (55.8) | **<.001** |
| Thal 4 | 364 (13.4) | 2 (0.5) | 9 (2.5) | 13 (3.6) | 50 (13.7) | 85 (23.4) | 110 (30.2) | 95 (26.1) | **<.001** |
| Thal 3 | 342 (12.6) | 10 (2.9) | 36 (10.5) | 45 (13.2) | 96 (28.1) | 95 (27.8) | 38 (11.1) | 22 (6.4) | **<.001** |
| Thal 2 | 226 (8.3) | 34 (15) | 38 (16.8) | 63 (27.9) | 50 (22.1) | 31 (13.7) | 5 (2.2) | 5 (2.2) | **<.001** |
| Thal 1 | 366 (13.5) | 56 (15.3) | 58 (15.8) | 75 (20.5) | 117 (32) | 57 (15.6) | 3 (0.8) | 0 (0) | **<.001** |
| Thal 0 | 1318 (48.6) | 438 (33.2) | 258 (19.6) | 230 (17.5) | 286 (21.7) | 94 (7.1) | 8 (0.6) | 4 (0.3) | **<.001** |
| **Female sex, No. (%)** | | | | | | | | | |
| Thal 5 | 73 (76.8) | 0 (0) | 1 (25) | NA | 3 (75) | 6 (66.7) | 19 (79.2) | 44 (83) | .06 |
| Thal 4 | 237 (65.1) | 1 (50) | 3 (33.3) | 5 (38.5) | 24 (48) | 54 (63.5) | 81 (73.6) | 69 (72.6) | **.02** |
| Thal 3 | 191 (55.8) | 5 (50) | 14 (38.9) | 26 (57.8) | 43 (44.8) | 62 (65.3) | 22 (57.9) | 19 (86.4) | **.008** |
| Thal 2 | 115 (50.9) | 18 (52.9) | 16 (42.1) | 38 (60.3) | 24 (48) | 14 (45.2) | 2 (40) | 3 (60) | .17 |
| Thal 1 | 175 (47.8) | 24 (42.9) | 28 (48.3) | 24 (32) | 72 (61.5) | 26 (45.6) | 1 (33.3) | NA | **.004** |
| Thal 0 | 549 (41.7) | 180 (41.1) | 90 (34.9) | 103 (44.8) | 120 (42) | 51 (54.3) | 3 (37.5) | 2 (50) | **.02** |
| **Age at death, mean (SD), y** | | | | | | | | | |
| Thal 5 | 83 (10.21) | 87.8 (NA) | 61.6 (18.05) | NA | 80 (10.01) | 87.3 (6.7) | 84 (9.07) | 83.5 (8.95) | >.99 |
| Thal 4 | 83.9 (9.23) | 73.6 (22.7) | 72 (10.19) | 77.1 (11.13) | 82.6 (9.8) | 84 (7.98) | 85.1 (8.82) | 85.3 (8.68) | >.99 |
| Thal 3 | 80.3 (11.43) | 66 (11.57) | 70 (10.93) | 76.1 (11.24) | 81.3 (10.93) | 83.2 (9.47) | 85.1 (8.61) | 86.5 (9.38) | **.004** |
| Thal 2 | 73.7 (11.63) | 62.9 (11.5) | 66.5 (8.33) | 75.4 (9.05) | 78.6 (9.14) | 81 (10.17) | 85.4 (10.28) | 76.2 (16.48) | **<.001** |
| Thal 1 | 75.2 (12.15) | 63.4 (11.55) | 70.6 (11.28) | 74.8 (9.58) | 79 (10.55) | 83.4 (8.84) | 88 (7.81) | NA | **<.001** |
| Thal 0 | 67.4 (14.57) | 57 (13.26) | 64.9 (10.73) | 72.4 (10.55) | 76.3 (11.61) | 81.9 (9.84) | 83.1 (11.99) | 75.9 (2.97) | **<.001** |
| **CDR >0, No. (%)** | | | | | | | | | |
| Thal 5 | 80 (84.2) | 1 (100) | 1 (25) | NA | 2 (50) | 6 (66.7) | 21 (87.5) | 49 (92.5) | **.01** |
| Thal 4 | 240 (65.9) | 2 (100) | 0 (0) | 6 (46.2) | 17 (34) | 51 (60) | 82 (74.5) | 82 (86.3) | **<.001** |
| Thal 3 | 153 (44.7) | 2 (20) | 6 (16.7) | 16 (35.6) | 39 (40.6) | 46 (48.4) | 25 (65.8) | 19 (86.4) | **<.001** |
| Thal 2 | 50 (22.1) | 3 (8.8) | 3 (7.9) | 15 (23.8) | 10 (20) | 14 (45.2) | 2 (40) | 3 (60) | **.004** |
| Thal 1 | 112 (30.6) | 13 (23.2) | 11 (19) | 26 (34.7) | 34 (29.1) | 25 (43.9) | 3 (100) | NA | **.04** |
| Thal 0 | 289 (21.9) | 67 (15.3) | 53 (20.5) | 49 (21.3) | 74 (25.9) | 39 (41.5) | 5 (62.5) | 2 (50) | **<.001** |
| **CDR ≥1, No. (%)** | | | | | | | | | |
| Thal 5 | 76 (80) | 1 (100) | 1 (25) | NA | 2 (50) | 6 (66.7) | 20 (83.3) | 46 (86.8) | **.03** |
| Thal 4 | 207 (56.9) | 1 (50) | 0 (0) | 5 (38.5) | 14 (28) | 39 (45.9) | 71 (64.5) | 77 (81.1) | **<.001** |
| Thal 3 | 117 (34.2) | 2 (20) | 2 (5.6) | 7 (15.6) | 33 (34.4) | 37 (38.9) | 19 (50) | 17 (77.3) | **<.001** |
| Thal 2 | 29 (12.8) | 2 (5.9) | 2 (5.3) | 9 (14.3) | 5 (10) | 8 (25.8) | 0 (0) | 3 (60) | **.01** |
| Thal 1 | 71 (19.4) | 9 (16.1) | 5 (8.6) | 16 (21.3) | 20 (17.1) | 18 (31.6) | 3 (100) | NA | **.005** |
| Thal 0 | 180 (13.7) | 36 (8.2) | 28 (10.9) | 30 (13) | 48 (16.8) | 32 (34) | 4 (50) | 2 (50) | **<.001** |
| **Contributing coexisting pathology, No. (%)** | | | | | | | | | |
| Thal 5 | 35 (36.8) | 0 (0) | 1 (25) | NA | 1 (25) | 3 (33.3) | 6 (25) | 24 (45.3) | .26 |
| Thal 4 | 133 (36.5) | 1 (50) | 1 (11.1) | 5 (38.5) | 12 (24) | 27 (31.8) | 40 (36.4) | 47 (49.5) | **.01** |
| Thal 3 | 80 (23.4) | 0 (0) | 10 (27.8) | 10 (22.2) | 15 (15.6) | 29 (30.5) | 8 (21.1) | 8 (36.4) | .09 |
| Thal 2 | 34 (15) | 2 (5.9) | 6 (15.8) | 11 (17.5) | 8 (16) | 7 (22.6) | 0 (0) | 0 (0) | .21 |
| Thal 1 | 68 (18.6) | 6 (10.7) | 10 (17.2) | 17 (22.7) | 16 (13.7) | 17 (29.8) | 2 (66.7) | NA | **.04** |
| Thal 0 | 191 (14.5) | 37 (8.4) | 38 (14.7) | 46 (20) | 39 (13.6) | 26 (27.7) | 4 (50) | 1 (25) | **<.001** |
| **APOE ε4 carrier, No. (%)** | | | | | | | | | |
| Thal 5 | 24 (25.3) | 0 (0) | 2 (50) | NA | 0 (0) | 2 (22.2) | 6 (25) | 14 (26.4) | >.99 |
| Thal 4 | 89 (24.5) | 0 (0) | 1 (11.1) | 3 (23.1) | 12 (24) | 23 (27.1) | 28 (25.5) | 22 (23.2) | >.99 |
| Thal 3 | 78 (22.8) | 2 (20) | 7 (19.4) | 11 (24.4) | 26 (27.1) | 23 (24.2) | 5 (13.2) | 4 (18.2) | .22 |
| Thal 2 | 50 (22.1) | 12 (35.3) | 10 (26.3) | 12 (19) | 10 (20) | 6 (19.4) | 0 (0) | 0 (0) | .10 |
| Thal 1 | 72 (19.7) | 11 (19.6) | 10 (17.2) | 17 (22.7) | 27 (23.1) | 7 (12.3) | 0 (0) | NA | .21 |
| Thal 0 | 137 (10.4) | 47 (10.7) | 34 (13.2) | 18 (7.8) | 30 (10.5) | 8 (8.5) | 0 (0) | 0 (0) | .17 |

Data are presented as mean (SD) or No. (%). In the first section, percentages in the Total column are based on all participants included in the table, whereas percentages in the Braak stage columns are calculated across each Thal phase row. Except for APOE ε4 carrier status, percentages in the remaining sections are calculated within the corresponding combination of Thal phase and Braak stage. CDR >0 includes scores of 0.5, 1, 2, or 3; CDR ≥1 includes scores of 1, 2, or 3. CDR ≥1 is reported to show that the gradient across Thal phase and Braak stage is not driven by the inclusion of very mild impairment (CDR 0.5); this threshold was descriptive and was not used in any inferential analysis. In the first section, 0 (0) indicates that no participant occupied that combination of Thal phase and Braak stage. In all later sections, 0 (0) indicates that the cell contained participants but none with the characteristic, whereas NA indicates that the cell contained no participants, so the proportion or mean is undefined. NA in the Age at death section also indicates that the SD could not be estimated because only 1 participant was present.P values are descriptive, unadjusted, 2-sided overall comparisons across Braak stages within each Thal phase row. No adjustment was made for multiple comparisons; given the number of such comparisons they are interpreted as descriptive only, and no threshold for statistical significance was applied. P values are reported to 2 decimal places unless P < .01, in which case 3 decimal places are used; values above .99 are reported as >.99. APOE ε4 carrier status was defined as ε2/ε4, ε3/ε4, or ε4/ε4. APOE ε4 counts reflect participants with available genotyping; percentages use all participants in the corresponding pathology stratum as the denominator. Genotyping was available for 1518 participants.

**Abbreviations:** APOE, apolipoprotein E; CDR, Clinical Dementia Rating; NA, not applicable; SD, standard deviation.

#### eTable 2. Demographic, Clinical, and Neuropathologic Characteristics by Braak Neurofibrillary Tangle Stage and CERAD Neuritic Plaque Score (n = 2711)

| Variable | Total | Braak stage | | | | | | | P value |
|---|---|---|---|---|---|---|---|---|---|
| | | 0 | I | II | III | IV | V | VI | |
| **CERAD neuritic plaque score, No. (%)** | | | | | | | | | |
| Frequent | 277 (10.2) | 2 (0.7) | 2 (0.7) | 2 (0.7) | 22 (7.9) | 38 (13.7) | 100 (36.1) | 111 (40.1) | **<.001** |
| Moderate | 393 (14.5) | 5 (1.3) | 25 (6.4) | 34 (8.7) | 82 (20.9) | 119 (30.3) | 71 (18.1) | 57 (14.5) | **<.001** |
| Sparse | 380 (14) | 27 (7.1) | 48 (12.6) | 91 (23.9) | 118 (31.1) | 82 (21.6) | 7 (1.8) | 7 (1.8) | **<.001** |
| None | 1661 (61.3) | 507 (30.5) | 328 (19.7) | 299 (18) | 381 (22.9) | 132 (7.9) | 10 (0.6) | 4 (0.2) | **<.001** |
| **Female sex, No. (%)** | | | | | | | | | |
| Frequent | 195 (70.4) | 1 (50) | 0 (0) | 2 (100) | 12 (54.5) | 19 (50) | 73 (73) | 88 (79.3) | **.01** |
| Moderate | 243 (61.8) | 2 (40) | 12 (48) | 12 (35.3) | 42 (51.2) | 82 (68.9) | 52 (73.2) | 41 (71.9) | **.003** |
| Sparse | 186 (48.9) | 11 (40.7) | 20 (41.7) | 47 (51.6) | 59 (50) | 43 (52.4) | 0 (0) | 6 (85.7) | **.03** |
| None | 716 (43.1) | 214 (42.2) | 120 (36.6) | 135 (45.2) | 173 (45.4) | 69 (52.3) | 3 (30) | 2 (50) | **.02** |
| **Age at death, mean (SD), y** | | | | | | | | | |
| Frequent | 84 (8.85) | 85.6 (3.18) | 63.6 (0.14) | 82.2 (1.34) | 81.1 (9.87) | 84.6 (8.85) | 84.5 (8.04) | 84.3 (9.14) | .62 |
| Moderate | 82.5 (9.98) | 74.8 (14.14) | 71.7 (10.19) | 76.8 (9.81) | 83 (9.73) | 83.4 (8.51) | 86.2 (9.16) | 84.4 (9.3) | .29 |
| Sparse | 78.5 (11.23) | 67.2 (11.22) | 69.3 (11.33) | 77.3 (8.6) | 80.7 (10) | 84.3 (8.99) | 83.6 (12.5) | 90.8 (8.27) | **<.001** |
| None | 68.3 (14.22) | 57.6 (13.02) | 65.7 (10.79) | 72.4 (10.6) | 76.5 (11.42) | 81.3 (9.78) | 81 (11.67) | 75.9 (2.97) | **<.001** |
| **CDR >0, No. (%)** | | | | | | | | | |
| Frequent | 211 (76.2) | 1 (50) | 1 (50) | 1 (50) | 6 (27.3) | 22 (57.9) | 80 (80) | 100 (90.1) | **<.001** |
| Moderate | 216 (55) | 2 (40) | 2 (8) | 10 (29.4) | 38 (46.3) | 71 (59.7) | 45 (63.4) | 48 (84.2) | **<.001** |
| Sparse | 129 (33.9) | 6 (22.2) | 8 (16.7) | 36 (39.6) | 31 (26.3) | 37 (45.1) | 6 (85.7) | 5 (71.4) | **.01** |
| None | 368 (22.2) | 79 (15.6) | 63 (19.2) | 65 (21.7) | 101 (26.5) | 51 (38.6) | 7 (70) | 2 (50) | **<.001** |
| **CDR ≥1, No. (%)** | | | | | | | | | |
| Frequent | 191 (69) | 1 (50) | 1 (50) | 0 (0) | 5 (22.7) | 18 (47.4) | 72 (72) | 94 (84.7) | **<.001** |
| Moderate | 179 (45.5) | 2 (40) | 1 (4) | 8 (23.5) | 32 (39) | 57 (47.9) | 35 (49.3) | 44 (77.2) | **<.001** |
| Sparse | 88 (23.2) | 5 (18.5) | 2 (4.2) | 22 (24.2) | 22 (18.6) | 27 (32.9) | 5 (71.4) | 5 (71.4) | **.003** |
| None | 222 (13.4) | 43 (8.5) | 34 (10.4) | 37 (12.4) | 63 (16.5) | 38 (28.8) | 5 (50) | 2 (50) | **<.001** |
| **Contributing coexisting pathology, No. (%)** | | | | | | | | | |
| Frequent | 103 (37.2) | 0 (0) | 1 (50) | 0 (0) | 5 (22.7) | 11 (28.9) | 32 (32) | 54 (48.6) | **.02** |
| Moderate | 115 (29.3) | 1 (20) | 3 (12) | 12 (35.3) | 19 (23.2) | 36 (30.3) | 20 (28.2) | 24 (42.1) | .05 |
| Sparse | 80 (21.1) | 4 (14.8) | 12 (25) | 24 (26.4) | 11 (9.3) | 25 (30.5) | 3 (42.9) | 1 (14.3) | **.003** |
| None | 243 (14.6) | 41 (8.1) | 50 (15.2) | 53 (17.7) | 56 (14.7) | 37 (28) | 5 (50) | 1 (25) | **<.001** |
| **APOE ε4 carrier, No. (%)** | | | | | | | | | |
| Frequent | 68 (24.5) | 0 (0) | 2 (100) | 0 (0) | 4 (18.2) | 11 (28.9) | 24 (24) | 27 (24.3) | .10 |
| Moderate | 84 (21.4) | 0 (0) | 5 (20) | 10 (29.4) | 18 (22) | 25 (21) | 13 (18.3) | 13 (22.8) | .35 |
| Sparse | 80 (21.1) | 7 (25.9) | 7 (14.6) | 16 (17.6) | 30 (25.4) | 19 (23.2) | 1 (14.3) | 0 (0) | .30 |
| None | 218 (13.1) | 65 (12.8) | 50 (15.2) | 35 (11.7) | 53 (13.9) | 14 (10.6) | 1 (10) | 0 (0) | .28 |

Data are presented as mean (SD) or No. (%). In the first section, percentages in the Total column are based on all participants included in the table, whereas percentages in the Braak stage columns are calculated across each CERAD neuritic plaque score row. Except for APOE ε4 carrier status, percentages in the remaining sections are calculated within the corresponding combination of CERAD neuritic plaque score and Braak stage. CDR >0 includes scores of 0.5, 1, 2, or 3; CDR ≥1 includes scores of 1, 2, or 3. CDR ≥1 is reported to show that the gradient across CERAD neuritic plaque score and Braak stage is not driven by the inclusion of very mild impairment (CDR 0.5); this threshold was descriptive and was not used in any inferential analysis. In the first section, 0 (0) indicates that no participant occupied that combination of CERAD neuritic plaque score and Braak stage. In all later sections, 0 (0) indicates that the cell contained participants but none with the characteristic, whereas NA indicates that the cell contained no participants, so the proportion or mean is undefined. NA in the Age at death section also indicates that the SD could not be estimated because only 1 participant was present.

P values are descriptive, unadjusted, 2-sided overall comparisons across Braak stages within each CERAD neuritic plaque score row. No adjustment was made for multiple comparisons; given the number of such comparisons they are interpreted as descriptive only, and no threshold for statistical significance was applied. P values are reported to 2 decimal places unless P < .01, in which case 3 decimal places are used; values above .99 are reported as >.99.

APOE ε4 carrier status was defined as ε2/ε4, ε3/ε4, or ε4/ε4. APOE ε4 counts reflect participants with available genotyping; percentages use all participants in the corresponding pathology stratum as the denominator. Genotyping was available for 1518 participants.

**Abbreviations:** APOE, apolipoprotein E; CDR, Clinical Dementia Rating; CERAD, Consortium to Establish a Registry for Alzheimer Disease; NA, not applicable; SD, standard deviation.

#### eTable 3. Demographic, Clinical, and Neuropathologic Characteristics by Braak Neurofibrillary Tangle Stage and Thal Amyloid Phase Among Female Participants (n = 1340)

| | | Braak stage | | | | | | | |
|---|---|---|---|---|---|---|---|---|---|
| **Variable** | **Total** | **0** | **I** | **II** | **III** | **IV** | **V** | **VI** | **P value** |
| **Thal phase, No. (%)** | | | | | | | | | |
| Thal 5 | 73 (5.4) | 0 (0) | 1 (1.4) | 0 (0) | 3 (4.1) | 6 (8.2) | 19 (26) | 44 (60.3) | **<.001** |
| Thal 4 | 237 (17.7) | 1 (0.4) | 3 (1.3) | 5 (2.1) | 24 (10.1) | 54 (22.8) | 81 (34.2) | 69 (29.1) | **<.001** |
| Thal 3 | 191 (14.3) | 5 (2.6) | 14 (7.3) | 26 (13.6) | 43 (22.5) | 62 (32.5) | 22 (11.5) | 19 (9.9) | **<.001** |
| Thal 2 | 115 (8.6) | 18 (15.7) | 16 (13.9) | 38 (33) | 24 (20.9) | 14 (12.2) | 2 (1.7) | 3 (2.6) | **<.001** |
| Thal 1 | 175 (13.1) | 24 (13.7) | 28 (16) | 24 (13.7) | 72 (41.1) | 26 (14.9) | 1 (0.6) | 0 (0) | **<.001** |
| Thal 0 | 549 (41) | 180 (32.8) | 90 (16.4) | 103 (18.8) | 120 (21.9) | 51 (9.3) | 3 (0.5) | 2 (0.4) | **<.001** |
| **Age at death, mean (SD), y** | | | | | | | | | |
| Thal 5 | 83.6 (9.25) | NA | 68.6 (NA) | NA | 78.4 (11.59) | 87.8 (7.12) | 84.7 (8.84) | 83.3 (9.4) | >.99 |
| Thal 4 | 85.9 (8.68) | 57.5 (NA) | 80.6 (10.58) | 76.6 (14.89) | 85 (8.71) | 85.8 (7.35) | 86.7 (8.42) | 86.9 (8.52) | >.99 |
| Thal 3 | 82.1 (10.52) | 69.3 (11.73) | 70.1 (7.92) | 76.7 (9.3) | 83.5 (10.97) | 84.3 (8.79) | 87.6 (7.69) | 85.4 (9.65) | .07 |
| Thal 2 | 74.9 (11.25) | 64.8 (12.46) | 67.4 (6.96) | 76.1 (9.49) | 78.5 (9.29) | 83.7 (8.02) | 87 (4.6) | 83.5 (9.26) | **.004** |
| Thal 1 | 78.3 (11.38) | 64.7 (10.89) | 74.6 (11.02) | 78.1 (8.86) | 81.6 (9.57) | 85.9 (6.71) | 81.9 (NA) | NA | **<.001** |
| Thal 0 | 69.5 (15.16) | 58.2 (14.38) | 66.1 (11.16) | 74.5 (10.16) | 77.5 (11.66) | 85 (9.08) | 79.9 (6.42) | 75.8 (5.02) | **<.001** |
| **CDR >0, No. (%)** | | | | | | | | | |
| Thal 5 | 67 (91.8) | NA | 0 (0) | NA | 2 (66.7) | 4 (66.7) | 19 (100) | 42 (95.5) | .13 |
| Thal 4 | 165 (69.6) | 1 (100) | 0 (0) | 4 (80) | 10 (41.7) | 30 (55.6) | 62 (76.5) | 58 (84.1) | **.006** |
| Thal 3 | 92 (48.2) | 2 (40) | 2 (14.3) | 7 (26.9) | 16 (37.2) | 35 (56.5) | 14 (63.6) | 16 (84.2) | **.004** |
| Thal 2 | 24 (20.9) | 0 (0) | 1 (6.2) | 9 (23.7) | 3 (12.5) | 8 (57.1) | 1 (50) | 2 (66.7) | **.003** |
| Thal 1 | 63 (36) | 7 (29.2) | 6 (21.4) | 9 (37.5) | 25 (34.7) | 15 (57.7) | 1 (100) | NA | **.04** |
| Thal 0 | 150 (27.3) | 38 (21.1) | 21 (23.3) | 30 (29.1) | 32 (26.7) | 27 (52.9) | 1 (33.3) | 1 (50) | **<.001** |
| **CDR ≥1, No. (%)** | | | | | | | | | |
| Thal 5 | 63 (86.3) | NA | 0 (0) | NA | 2 (66.7) | 4 (66.7) | 18 (94.7) | 39 (88.6) | .29 |
| Thal 4 | 147 (62) | 0 (0) | 0 (0) | 3 (60) | 8 (33.3) | 28 (51.9) | 53 (65.4) | 55 (79.7) | **.004** |
| Thal 3 | 75 (39.3) | 2 (40) | 1 (7.1) | 3 (11.5) | 16 (37.2) | 29 (46.8) | 9 (40.9) | 15 (78.9) | **<.001** |
| Thal 2 | 15 (13) | 0 (0) | 0 (0) | 6 (15.8) | 2 (8.3) | 5 (35.7) | 0 (0) | 2 (66.7) | **.03** |
| Thal 1 | 44 (25.1) | 4 (16.7) | 5 (17.9) | 5 (20.8) | 15 (20.8) | 14 (53.8) | 1 (100) | NA | **.002** |
| Thal 0 | 99 (18) | 22 (12.2) | 9 (10) | 19 (18.4) | 23 (19.2) | 24 (47.1) | 1 (33.3) | 1 (50) | **<.001** |
| **Contributing coexisting pathology, No. (%)** | | | | | | | | | |
| Thal 5 | 29 (39.7) | NA | 0 (0) | NA | 1 (33.3) | 2 (33.3) | 5 (26.3) | 21 (47.7) | .34 |
| Thal 4 | 88 (37.1) | 0 (0) | 1 (33.3) | 2 (40) | 6 (25) | 18 (33.3) | 30 (37) | 31 (44.9) | .22 |
| Thal 3 | 44 (23) | 0 (0) | 3 (21.4) | 5 (19.2) | 5 (11.6) | 19 (30.6) | 5 (22.7) | 7 (36.8) | .11 |
| Thal 2 | 13 (11.3) | 2 (11.1) | 0 (0) | 7 (18.4) | 2 (8.3) | 2 (14.3) | 0 (0) | 0 (0) | .26 |
| Thal 1 | 34 (19.4) | 4 (16.7) | 5 (17.9) | 6 (25) | 9 (12.5) | 9 (34.6) | 1 (100) | NA | .09 |
| Thal 0 | 89 (16.2) | 18 (10) | 13 (14.4) | 26 (25.2) | 15 (12.5) | 16 (31.4) | 1 (33.3) | 0 (0) | **.006** |
| **APOE ε4 carrier, No. (%)** | | | | | | | | | |
| Thal 5 | 18 (24.7) | NA | 0 (0) | NA | 0 (0) | 2 (33.3) | 6 (31.6) | 10 (22.7) | .67 |
| Thal 4 | 60 (25.3) | 0 (0) | 0 (0) | 1 (20) | 4 (16.7) | 17 (31.5) | 21 (25.9) | 17 (24.6) | .37 |
| Thal 3 | 40 (20.9) | 0 (0) | 2 (14.3) | 6 (23.1) | 11 (25.6) | 15 (24.2) | 2 (9.1) | 4 (21.1) | .27 |
| Thal 2 | 21 (18.3) | 7 (38.9) | 5 (31.2) | 6 (15.8) | 3 (12.5) | 0 (0) | 0 (0) | 0 (0) | .05 |
| Thal 1 | 27 (15.4) | 5 (20.8) | 4 (14.3) | 3 (12.5) | 11 (15.3) | 4 (15.4) | 0 (0) | NA | .65 |
| Thal 0 | 58 (10.6) | 20 (11.1) | 16 (17.8) | 9 (8.7) | 10 (8.3) | 3 (5.9) | 0 (0) | 0 (0) | .04 |

Data are presented as mean (SD) or No. (%). In the first section, percentages in the Total column are based on all participants included in the table, whereas percentages in the Braak stage columns are calculated across each Thal phase row. Except for APOE ε4 carrier status, percentages in the remaining sections are calculated within the corresponding combination of Thal phase and Braak stage. CDR >0 includes scores of 0.5, 1, 2, or 3; CDR ≥1 includes scores of 1, 2, or 3. CDR ≥1 is reported to show that the gradient across Thal phase and Braak stage is not driven by the inclusion of very mild impairment (CDR 0.5); this threshold was descriptive and was not used in any inferential analysis. In the first section, 0 (0) indicates that no participant occupied that combination of Thal phase and Braak stage. In all later sections, 0 (0) indicates that the cell contained participants but none with the characteristic, whereas NA indicates that the cell contained no participants, so the proportion or mean is undefined. NA in the Age at death section also indicates that the SD could not be estimated because only 1 participant was present.

P values are descriptive, unadjusted, 2-sided overall comparisons across Braak stages within each Thal phase row. No adjustment was made for multiple comparisons; given the number of such comparisons they are interpreted as descriptive only, and no threshold for statistical significance was applied. P values are reported to 2 decimal places unless P < .01, in which case 3 decimal places are used; values above .99 are reported as >.99. APOE ε4 carrier status was defined as ε2/ε4, ε3/ε4, or ε4/ε4. APOE ε4 counts reflect participants with available genotyping; percentages use all participants in the corresponding pathology stratum as the denominator. Genotyping was available for 731 participants.

**Abbreviations:** APOE, apolipoprotein E; CDR, Clinical Dementia Rating; NA, not applicable; SD, standard deviation.

#### eTable 4. Demographic, Clinical, and Neuropathologic Characteristics by Braak Neurofibrillary Tangle Stage and Thal Amyloid Phase Among Male Participants (n = 1371)

| | | Braak stage | | | | | | | |
|---|---|---|---|---|---|---|---|---|---|
| **Variable** | **Total** | **0** | **I** | **II** | **III** | **IV** | **V** | **VI** | **P value** |
| **Thal phase, No. (%)** | | | | | | | | | |
| Thal 5 | 22 (1.6) | 1 (4.5) | 3 (13.6) | 0 (0) | 1 (4.5) | 3 (13.6) | 5 (22.7) | 9 (40.9) | .006 |
| Thal 4 | 127 (9.3) | 1 (0.8) | 6 (4.7) | 8 (6.3) | 26 (20.5) | 31 (24.4) | 29 (22.8) | 26 (20.5) | <.001 |
| Thal 3 | 151 (11) | 5 (3.3) | 22 (14.6) | 19 (12.6) | 53 (35.1) | 33 (21.9) | 16 (10.6) | 3 (2) | <.001 |
| Thal 2 | 111 (8.1) | 16 (14.4) | 22 (19.8) | 25 (22.5) | 26 (23.4) | 17 (15.3) | 3 (2.7) | 2 (1.8) | <.001 |
| Thal 1 | 191 (13.9) | 32 (16.8) | 30 (15.7) | 51 (26.7) | 45 (23.6) | 31 (16.2) | 2 (1) | 0 (0) | <.001 |
| Thal 0 | 769 (56.1) | 258 (33.6) | 168 (21.8) | 127 (16.5) | 166 (21.6) | 43 (5.6) | 5 (0.7) | 2 (0.3) | <.001 |
| **Age at death, mean (SD), y** | | | | | | | | | |
| Thal 5 | 80.7 (12.89) | 87.8 (NA) | 59.2 (21.35) | NA | 84.9 (NA) | 86.1 (7.05) | 81.6 (10.6) | 84.4 (6.71) | >.99 |
| Thal 4 | 80 (8.98) | 89.6 (NA) | 67.7 (7.41) | 77.4 (9.24) | 80.5 (10.42) | 80.9 (8.2) | 80.4 (8.36) | 81 (7.73) | >.99 |
| Thal 3 | 77.9 (12.11) | 62.6 (11.65) | 70 (12.65) | 75.3 (13.69) | 79.5 (10.67) | 81.1 (10.48) | 81.6 (8.85) | 93.4 (2) | .04 |
| Thal 2 | 72.5 (11.94) | 60.7 (10.28) | 65.8 (9.3) | 74.4 (8.43) | 78.6 (9.19) | 78.9 (11.43) | 84.3 (14) | 65.2 (22.63) | <.001 |
| Thal 1 | 72.4 (12.16) | 62.5 (12.1) | 66.9 (10.38) | 73.3 (9.59) | 75 (10.89) | 81.4 (9.94) | 91 (8.13) | NA | <.001 |
| Thal 0 | 66 (13.96) | 56.2 (12.38) | 64.3 (10.48) | 70.8 (10.6) | 75.4 (11.54) | 78.1 (9.47) | 85 (14.79) | 75.9 (1.13) | <.001 |
| **CDR >0, No. (%)** | | | | | | | | | |
| Thal 5 | 13 (59.1) | 1 (100) | 1 (33.3) | NA | 0 (0) | 2 (66.7) | 2 (40) | 7 (77.8) | .42 |
| Thal 4 | 75 (59.1) | 1 (100) | 0 (0) | 2 (25) | 7 (26.9) | 21 (67.7) | 20 (69) | 24 (92.3) | .001 |
| Thal 3 | 61 (40.4) | 0 (0) | 4 (18.2) | 9 (47.4) | 23 (43.4) | 11 (33.3) | 11 (68.8) | 3 (100) | .04 |
| Thal 2 | 26 (23.4) | 3 (18.8) | 2 (9.1) | 6 (24) | 7 (26.9) | 6 (35.3) | 1 (33.3) | 1 (50) | .18 |
| Thal 1 | 49 (25.7) | 6 (18.8) | 5 (16.7) | 17 (33.3) | 9 (20) | 10 (32.3) | 2 (100) | NA | .11 |
| Thal 0 | 139 (18.1) | 29 (11.2) | 32 (19) | 19 (15) | 42 (25.3) | 12 (27.9) | 4 (80) | 1 (50) | .003 |
| **CDR ≥1, No. (%)** | | | | | | | | | |
| Thal 5 | 13 (59.1) | 1 (100) | 1 (33.3) | NA | 0 (0) | 2 (66.7) | 2 (40) | 7 (77.8) | .42 |
| Thal 4 | 60 (47.2) | 1 (100) | 0 (0) | 2 (25) | 6 (23.1) | 11 (35.5) | 18 (62.1) | 22 (84.6) | <.001 |
| Thal 3 | 42 (27.8) | 0 (0) | 1 (4.5) | 4 (21.1) | 17 (32.1) | 8 (24.2) | 10 (62.5) | 2 (66.7) | .005 |
| Thal 2 | 14 (12.6) | 2 (12.5) | 2 (9.1) | 3 (12) | 3 (11.5) | 3 (17.6) | 0 (0) | 1 (50) | .60 |
| Thal 1 | 27 (14.1) | 5 (15.6) | 0 (0) | 11 (21.6) | 5 (11.1) | 4 (12.9) | 2 (100) | NA | .01 |
| Thal 0 | 81 (10.5) | 14 (5.4) | 19 (11.3) | 11 (8.7) | 25 (15.1) | 8 (18.6) | 3 (60) | 1 (50) | .004 |
| **Contributing coexisting pathology, No. (%)** | | | | | | | | | |
| Thal 5 | 6 (27.3) | 0 (0) | 1 (33.3) | NA | 0 (0) | 1 (33.3) | 1 (20) | 3 (33.3) | .97 |
| Thal 4 | 45 (35.4) | 1 (100) | 0 (0) | 3 (37.5) | 6 (23.1) | 9 (29) | 10 (34.5) | 16 (61.5) | .01 |
| Thal 3 | 36 (23.8) | 0 (0) | 7 (31.8) | 5 (26.3) | 10 (18.9) | 10 (30.3) | 3 (18.8) | 1 (33.3) | .47 |
| Thal 2 | 21 (18.9) | 0 (0) | 6 (27.3) | 4 (16) | 6 (23.1) | 5 (29.4) | 0 (0) | 0 (0) | .11 |
| Thal 1 | 34 (17.8) | 2 (6.2) | 5 (16.7) | 11 (21.6) | 7 (15.6) | 8 (25.8) | 1 (50) | NA | .14 |
| Thal 0 | 102 (13.3) | 19 (7.4) | 25 (14.9) | 20 (15.7) | 24 (14.5) | 10 (23.3) | 3 (60) | 1 (50) | .007 |
| **APOE ε4 carrier, No. (%)** | | | | | | | | | |
| Thal 5 | 6 (27.3) | 0 (0) | 2 (66.7) | NA | 0 (0) | 0 (0) | 0 (0) | 4 (44.4) | .38 |
| Thal 4 | 29 (22.8) | 0 (0) | 1 (16.7) | 2 (25) | 8 (30.8) | 6 (19.4) | 7 (24.1) | 5 (19.2) | .46 |
| Thal 3 | 38 (25.2) | 2 (40) | 5 (22.7) | 5 (26.3) | 15 (28.3) | 8 (24.2) | 3 (18.8) | 0 (0) | .72 |
| Thal 2 | 29 (26.1) | 5 (31.2) | 5 (22.7) | 6 (24) | 7 (26.9) | 6 (35.3) | 0 (0) | 0 (0) | .56 |
| Thal 1 | 45 (23.6) | 6 (18.8) | 6 (20) | 14 (27.5) | 16 (35.6) | 3 (9.7) | 0 (0) | NA | .09 |
| Thal 0 | 79 (10.3) | 27 (10.5) | 18 (10.7) | 9 (7.1) | 20 (12) | 5 (11.6) | 0 (0) | 0 (0) | .30 |

Data are presented as mean (SD) or No. (%). In the first section, percentages in the Total column are based on all participants included in the table, whereas percentages in the Braak stage columns are calculated across each Thal phase row. Except for APOE ε4 carrier status, percentages in the remaining sections are calculated within the corresponding combination of Thal phase and Braak stage. CDR >0 includes scores of 0.5, 1, 2, or 3; CDR ≥1 includes scores of 1, 2, or 3. CDR ≥1 is reported to show that the gradient across Thal phase and Braak stage is not driven by the inclusion of very mild impairment (CDR 0.5); this threshold was descriptive and was not used in any inferential analysis. In the first section, 0 (0) indicates that no participant occupied that combination of Thal phase and Braak stage. In all later sections, 0 (0) indicates that the cell contained participants but none with the characteristic, whereas NA indicates that the cell contained no participants, so the proportion or mean is undefined. NA in the Age at death section also indicates that the SD could not be estimated because only 1 participant was present.

P values are descriptive, unadjusted, 2-sided overall comparisons across Braak stages within each Thal phase row. No adjustment was made for multiple comparisons; given the number of such comparisons, they are interpreted as descriptive only, and no threshold for statistical significance was applied. P values are reported to 2 decimal places unless P < .01, in which case 3 decimal places are used; values above .99 are reported as >.99. APOE ε4 carrier status was defined as ε2/ε4, ε3/ε4, or ε4/ε4. APOE ε4 counts reflect participants with available genotyping; percentages use all participants in the corresponding pathology stratum as the denominator. Genotyping was available for 787 participants.

**Abbreviations:** APOE, apolipoprotein E; CDR, Clinical Dementia Rating; NA, not applicable; SD, standard deviation.

### eTable 5. Demographic, Clinical, and Neuropathologic Characteristics by Braak Neurofibrillary Tangle Stage and Thal Amyloid Phase Among Participants Without Contributing Coexisting Pathology (n = 2170)

| | | Braak stage | | | | | | | |
|---|---|---|---|---|---|---|---|---|---|
| **Variable** | **Total** | **0** | **I** | **II** | **III** | **IV** | **V** | **VI** | **P value** |
| **Thal phase, No. (%)** | | | | | | | | | |
| Thal 5 | 60 (2.8) | 1 (1.7) | 3 (5) | 0 (0) | 3 (5) | 6 (10) | 18 (30) | 29 (48.3) | **<.001** |
| Thal 4 | 231 (10.6) | 1 (0.4) | 8 (3.5) | 8 (3.5) | 38 (16.5) | 58 (25.1) | 70 (30.3) | 48 (20.8) | **<.001** |
| Thal 3 | 262 (12.1) | 10 (3.8) | 26 (9.9) | 35 (13.4) | 81 (30.9) | 66 (25.2) | 30 (11.5) | 14 (5.3) | **<.001** |
| Thal 2 | 192 (8.8) | 32 (16.7) | 32 (16.7) | 52 (27.1) | 42 (21.9) | 24 (12.5) | 5 (2.6) | 5 (2.6) | **<.001** |
| Thal 1 | 298 (13.7) | 50 (16.8) | 48 (16.1) | 58 (19.5) | 101 (33.9) | 40 (13.4) | 1 (0.3) | 0 (0) | **<.001** |
| Thal 0 | 1127 (51.9) | 401 (35.6) | 220 (19.5) | 184 (16.3) | 247 (21.9) | 68 (6) | 4 (0.4) | 3 (0.3) | **<.001** |
| **Female sex, No. (%)** | | | | | | | | | |
| Thal 5 | 44 (73.3) | 0 (0) | 1 (33.3) | NA | 2 (66.7) | 4 (66.7) | 14 (77.8) | 23 (79.3) | .36 |
| Thal 4 | 149 (64.5) | 1 (100) | 2 (25) | 3 (37.5) | 18 (47.4) | 36 (62.1) | 51 (72.9) | 38 (79.2) | **.04** |
| Thal 3 | 147 (56.1) | 5 (50) | 11 (42.3) | 21 (60) | 38 (46.9) | 43 (65.2) | 17 (56.7) | 12 (85.7) | .05 |
| Thal 2 | 102 (53.1) | 16 (50) | 16 (50) | 31 (59.6) | 22 (52.4) | 12 (50) | 2 (40) | 3 (60) | .42 |
| Thal 1 | 141 (47.3) | 20 (40) | 23 (47.9) | 18 (31) | 63 (62.4) | 17 (42.5) | 0 (0) | NA | **.003** |
| Thal 0 | 460 (40.8) | 162 (40.4) | 77 (35) | 77 (41.8) | 105 (42.5) | 35 (51.5) | 2 (50) | 2 (66.7) | .09 |
| **Age at death, mean (SD), y** | | | | | | | | | |
| Thal 5 | 82.6 (11.08) | 87.8 (NA) | 60.9 (22.04) | NA | 77.3 (10.28) | 87.2 (8.15) | 84.1 (8.62) | 83.3 (9.91) | >.99 |
| Thal 4 | 83 (9.57) | 57.5 (NA) | 71.3 (10.67) | 79.8 (7.84) | 81.9 (10.26) | 83.4 (7.83) | 83.7 (9.23) | 85.4 (9.65) | >.99 |
| Thal 3 | 79.3 (11.41) | 66 (11.57) | 70.2 (11.04) | 75.2 (12.11) | 80.1 (11.09) | 82 (8.24) | 85.3 (9.22) | 86.1 (9.73) | **.005** |
| Thal 2 | 73 (11.83) | 62.1 (11.43) | 66.3 (8.47) | 74.1 (8.79) | 78.5 (9.61) | 80.9 (10.12) | 85.4 (10.28) | 76.2 (16.48) | **<.001** |
| Thal 1 | 74.2 (12.16) | 62.9 (11.76) | 68.8 (11.03) | 73.6 (9.16) | 79.2 (10.14) | 82.2 (8.58) | 96.8 (NA) | NA | **<.001** |
| Thal 0 | 66.2 (14.74) | 56.2 (13.14) | 64.1 (10.93) | 71.8 (10.83) | 75.8 (11.56) | 81.1 (10.43) | 88.8 (15.15) | 75.6 (3.58) | **<.001** |
| **CDR >0, No. (%)** | | | | | | | | | |
| Thal 5 | 46 (76.7) | 1 (100) | 0 (0) | NA | 1 (33.3) | 4 (66.7) | 15 (83.3) | 25 (86.2) | **.01** |
| Thal 4 | 131 (56.7) | 1 (100) | 0 (0) | 3 (37.5) | 13 (34.2) | 31 (53.4) | 47 (67.1) | 36 (75) | **.004** |
| Thal 3 | 98 (37.4) | 2 (20) | 2 (7.7) | 10 (28.6) | 26 (32.1) | 28 (42.4) | 19 (63.3) | 11 (78.6) | **.003** |
| Thal 2 | 37 (19.3) | 3 (9.4) | 2 (6.2) | 9 (17.3) | 7 (16.7) | 11 (45.8) | 2 (40) | 3 (60) | **.002** |
| Thal 1 | 68 (22.8) | 8 (16) | 7 (14.6) | 14 (24.1) | 25 (24.8) | 13 (32.5) | 1 (100) | NA | .20 |
| Thal 0 | 175 (15.5) | 44 (11) | 31 (14.1) | 21 (11.4) | 54 (21.9) | 21 (30.9) | 3 (75) | 1 (33.3) | **<.001** |
| **CDR ≥1, No. (%)** | | | | | | | | | |
| Thal 5 | 43 (71.7) | 1 (100) | 0 (0) | NA | 1 (33.3) | 4 (66.7) | 14 (77.8) | 23 (79.3) | .03 |
| Thal 4 | 103 (44.6) | 0 (0) | 0 (0) | 2 (25) | 10 (26.3) | 20 (34.5) | 38 (54.3) | 33 (68.8) | **.001** |
| Thal 3 | 73 (27.9) | 2 (20) | 1 (3.8) | 3 (8.6) | 22 (27.2) | 21 (31.8) | 15 (50) | 9 (64.3) | **.006** |
| Thal 2 | 20 (10.4) | 2 (6.2) | 1 (3.1) | 6 (11.5) | 3 (7.1) | 5 (20.8) | 0 (0) | 3 (60) | **.004** |
| Thal 1 | 35 (11.7) | 5 (10) | 2 (4.2) | 6 (10.3) | 13 (12.9) | 8 (20) | 1 (100) | NA | .16 |
| Thal 0 | 94 (8.3) | 19 (4.7) | 15 (6.8) | 10 (5.4) | 32 (13) | 15 (22.1) | 2 (50) | 1 (33.3) | **<.001** |
| **APOE ε4 carrier, No. (%)** | | | | | | | | | |
| Thal 5 | 16 (26.7) | 0 (0) | 1 (33.3) | NA | 0 (0) | 1 (16.7) | 4 (22.2) | 10 (34.5) | .46 |
| Thal 4 | 60 (26) | 0 (0) | 1 (12.5) | 2 (25) | 10 (26.3) | 15 (25.9) | 18 (25.7) | 14 (29.2) | .64 |
| Thal 3 | 64 (24.4) | 2 (20) | 6 (23.1) | 11 (31.4) | 21 (25.9) | 17 (25.8) | 4 (13.3) | 3 (21.4) | .23 |
| Thal 2 | 41 (21.4) | 11 (34.4) | 9 (28.1) | 10 (19.2) | 8 (19) | 3 (12.5) | 0 (0) | 0 (0) | .12 |
| Thal 1 | 63 (21.1) | 10 (20) | 9 (18.8) | 15 (25.9) | 25 (24.8) | 4 (10) | 0 (0) | NA | .13 |
| Thal 0 | 122 (10.8) | 42 (10.5) | 31 (14.1) | 15 (8.2) | 26 (10.5) | 8 (11.8) | 0 (0) | 0 (0) | .14 |

Data are presented as mean (SD) or No. (%). In the first section, percentages in the Total column are based on all participants included in the table, whereas percentages in the Braak stage columns are calculated across each Thal phase row. Except for APOE ε4 carrier status, percentages in the remaining sections are calculated within the corresponding combination of Thal phase and Braak stage. CDR >0 includes scores of 0.5, 1, 2, or 3; CDR ≥1 includes scores of 1, 2, or 3. CDR ≥1 is reported to show that the gradient across Thal phase and Braak stage is not driven by the inclusion of very mild impairment (CDR 0.5); this threshold was descriptive and was not used in any inferential analysis. In the first section, 0 (0) indicates that no participant occupied that combination of Thal phase and Braak stage. In all later sections, 0 (0) indicates that the cell contained participants but none with the characteristic, whereas NA indicates that the cell contained no participants, so the proportion or mean is undefined. NA in the Age at death section also indicates that the SD could not be estimated because only 1 participant was present.

P values are descriptive, unadjusted, 2-sided overall comparisons across Braak stages within each Thal phase row. No adjustment was made for multiple comparisons; given the number of such comparisons they are interpreted as descriptive only, and no threshold for statistical significance was applied. P values are reported to 2 decimal places unless P < .01, in which case 3 decimal places are used; values above .99 are reported as >.99. APOE ε4 carrier status was defined as ε2/ε4, ε3/ε4, or ε4/ε4. APOE ε4 counts reflect participants with available genotyping; percentages use all participants in the corresponding pathology stratum as the denominator. Genotyping was available for 1265 participants.

**Abbreviations:** APOE, apolipoprotein E; CDR, Clinical Dementia Rating; NA, not applicable; SD, standard deviation.

### eTable 6. Coefficient Estimates from Candidate Multivariable Linear Regression Models of Clinical Dementia Rating Sum of Boxes

| **Variable** | ***m0*** | | ***m1*** | |
|---|---|---|---|---|
| Analytic No. / % | 1518 / 56 | | 2711 / 100 | |
| | **β (95% CI)** | **P value** | **β (95% CI)** | **P value** |
| Braak stage I | -0.59 (-1.384 to 0.199) | .14 | -0.9 (-1.560 to -0.239) | **.008** |
| Braak stage II | -0.49 (-1.355 to 0.369) | .26 | -0.62 (-1.311 to 0.068) | .08 |
| Braak stage III | 0.03 (-0.809 to 0.861) | .95 | -0.23 (-0.904 to 0.448) | .51 |
| Braak stage IV | 1.46 (0.476 to 2.449) | **.004** | 1.22 (0.407 to 2.039) | **.003** |
| Braak stage V | 3.91 (2.458 to 5.369) | **<.001** | 3.34 (2.212 to 4.466) | **<.001** |
| Braak stage VI | 6.74 (5.150 to 8.328) | **<.001** | 6.13 (4.932 to 7.324) | **<.001** |
| Contributing coexisting pathology | 4.16 (3.519 to 4.810) | **<.001** | 4.03 (3.538 to 4.517) | **<.001** |
| CERAD neuritic plaque score 1 | 0.62 (-0.309 to 1.555) | .19 | 0.57 (-0.207 to 1.343) | .15 |
| CERAD neuritic plaque score 2 | 0.83 (-0.429 to 2.094) | .20 | 1.48 (0.496 to 2.459) | **.003** |
| CERAD neuritic plaque score 3 | 1.97 (0.434 to 3.498) | **.01** | 2.38 (1.212 to 3.558) | **<.001** |
| Thal phase 1 | -0.48 (-1.237 to 0.285) | .22 | -0.17 (-0.831 to 0.484) | .61 |
| Thal phase 2 | -1.54 (-2.535 to -0.544) | **.002** | -1.34 (-2.163 to -0.520) | **<.001** |
| Thal phase 3 | -0.8 (-1.977 to 0.383) | .19 | -0.52 (-1.459 to 0.426) | .28 |
| Thal phase 4 | -0.75 (-2.157 to 0.658) | .30 | -0.3 (-1.386 to 0.794) | .59 |
| Thal phase 5 | 2.11 (0.281 to 3.947) | **.02** | 2.19 (0.735 to 3.642) | **.003** |
| Female sex | 0.54 (0.058 to 1.019) | **.03** | 0.79 (0.404 to 1.183) | **<.001** |
| Age at death, per 10-y increase | 0.05 (0.027 to 0.071) | **<.001** | 0.06 (0.045 to 0.079) | **<.001** |
| Education, per 1-y increase | -0.09 (-0.149 to -0.037) | **<.001** | - | - |
| APOE ε4 carrier | 0.3 (-0.228 to 0.833) | .26 | - | - |

Values are β coefficients (95% CIs) and represent adjusted differences in CDR-SB points. Age at death was modeled per 10-year increase, and education per 1-year increase. Binary comparisons are presence vs. absence of contributing coexisting pathology, APOE ε4 carrier vs. noncarrier, and female vs. male. A dash indicates that the predictor was not included in the model. Models used complete cases for the variables included, so the analytic No. varies across models and is reported in the first row of each model column.

**Abbreviations:** APOE, apolipoprotein E; CDR-SB, Clinical Dementia Rating Sum of Boxes; CERAD, Consortium to Establish a Registry for Alzheimer Disease; CI, confidence interval.

#### eTable 7. Exploratory Positive and Negative Predictive Values for a Plasma Phosphorylated Tau 217 Assay by Age Stratum, Using Tissue-Derived Frequency

| Age at death, y | Frequency, % | PPV, % (95% range) | | NPV, % (95% range) | |
|---|---|---|---|---|---|
| | | Unpenalized | Optimism-corrected | Unpenalized | Optimism-corrected |
| 23-50 | 0.0 | Not estimable | Not estimable | Not estimable | Not estimable |
| 51-60 | 2.0 | 12.2 (6.9-48.8) | 9.4 (6.0-20.9) | 99.8 (99.6-100.0) | 99.7 (99.5-99.8) |
| 61-70 | 5.5 | 28.4 (17.4-73.1) | 22.9 (15.4-43.0) | 99.4 (99.0-99.9) | 99.1 (98.7-99.5) |
| 71-80 | 14.5 | 53.6 (38.1-88.8) | 46.4 (34.7-68.7) | 98.4 (97.0-99.6) | 97.5 (96.2-98.6) |
| 81-90 | 36.5 | 79.7 (67.6-96.4) | 74.6 (64.3-88.1) | 94.7 (90.7-98.6) | 92.0 (88.2-95.5) |
| 91-111 | 45.9 | 85.2 (75.5-97.5) | 81.3 (72.7-91.6) | 92.4 (86.8-98.0) | 88.6 (83.5-93.6) |
| All ages | 19.6 | 62.4 (46.9-91.9) | 55.5 (43.3-75.9) | 97.7 (95.8-99.4) | 96.4 (94.6-98.1) |

Frequency is the observed age-specific frequency of combined biomarker-detectable amyloid and tau neuropathology, defined as Thal amyloid phase 3 to 5 together with Braak neurofibrillary tangle stage IV to VI, in the analytic cohort (n = 2711). Predictive values were computed by the Bayes formulas PPV = (Se × Prev)/(Se × Prev + (1 − Sp) × (1 − Prev)) and NPV = (Sp × (1 − Prev))/((1 − Se) × Prev + Sp × (1 − Prev)), enumerating all combinations of externally derived sensitivity (0.850-0.982) and specificity (0.745-0.986) in 0.001 increments. Values are medians with 2.5th to 97.5th percentiles of the resulting distributions. Optimism-corrected values were obtained after shrinking sensitivity and specificity 10% toward 0.5 (Se* = 0.5 + 0.9 × (Se − 0.5); Sp* analogous), because externally derived assay performance typically attenuates on transfer to a new population. At median sensitivity and specificity, PPV reaches 50% at a frequency of 12.8%, 75% at 30.6%, and 90% at 56.9%.

**Abbreviations:** NPV, negative predictive value; PPV, positive predictive value; Prev, frequency of the target condition; Se, sensitivity; Sp, specificity.
